# Timing HIV infection with nonlinear viral dynamics

**DOI:** 10.1101/2020.08.13.20174243

**Authors:** Daniel Reeves, Morgane Rolland, Bethany L Dearlove, Yifan Li, Merlin Robb, Joshua T Schiffer, Peter Gilbert, E Fabian Cardozo-Ojeda, Bryan Mayer

**Author notes:** These authors contributed equally (E Fabian Cardozo-Ojeda, Bryan Mayer).

## Abstract

In HIV prevention trials, precise identification of infection time is critical to quantify drug efficacy but difficult to estimate as trials may have relatively sparse visit schedules. The last negative visit does not guarantee a boundary on infection time because viral nucleic acid is not present in the blood during early infection. Here, we developed a framework that combines stochastic and deterministic within-host mathematical modeling of viral dynamics accounting for the early unobservable viral load phase until it reaches a high chronic set point. The infection time estimation is based on a population non-linear mixed effects (pNLME) framework that includes the with-in host modeling. We applied this framework to viral load data from the RV217 trial and found a parsimonious model capable of recapitulating the viral loads. When adding the stochastic and deterministic portion of the best model, the estimated infection time for the RV217 data had an average of 2 weeks between infecting exposure and first positive. We assessed the sensitivity of the infection time estimation by conducting *in silico* studies with varying viral load sampling schemes before and after infection. pNLME accurately estimates infection times for a daily sampling scheme and is fairly robust to sparser schemes. For a monthly sampling scheme before and after first positive bias increases to -7 days. For pragmatic trial design, we found sampling weekly before and monthly after first positive allows accurate pNLME estimation. Our estimates can be used in parallel with other approaches that rely on viral sequencing, and because the model is mechanistic, it is primed for future application to infection timing for specific interventions.

## Introduction

A key challenge for HIV prevention trials is to identify the timing of the exposure that ultimately led to breakthrough infection. Estimation of infection time subsequently allows inference of the concentration of the protective agent at exposure, which is critical to understanding why HIV acquisition was not prevented. Early infection is difficult to study in practice; even if prospective sampling were available, HIV RNA is not detectable in blood during early HIV infection and participants cannot necessarily point to potential recent exposure events with accuracy. Therefore, to estimate time of infection, a model or inference technique is required.

Estimation techniques have been described previously. Several use viral sequence data and evolutionary models to trace time back to the founder sequence^1-3^. Others use viral load data prior to viral peak and retrace using log-linear regression (average or maximum upslope)^3^. Others apply diagnostic ‘window times’ that leverage Fiebig staging^4^ and prior knowledge of eclipse phase duration^5,6^, where eclipse phase is defined as the period of time between HIV acquisition and first detectable viral load. Finally, combinations of these approaches have been organized into a statistical framework^7^.

Here, we introduce an approach applying such viral dynamics models with statistical inference on viral load data. Model development was achieved through fitting to longitudinally sampled viral loads from 46 participants in the RV217 study^8^. Population nonlinear mixed-effects (pNLME) modeling was used to determine the optimal model parameters for each individual, given a population distribution. We tested 30 candidate models informed by previous viral load modeling, and the best model was selected by parsimony. The very early moments of HIV infection are thought to be stochastic^9^, and have been modeled with stochastic viral dynamics^10,11^. Therefore, we used our best model in a stochastic formulation to simulate early HIV dynamics, allowing for fluctuations and extinction by chance. Together, the stochastic and deterministic models provide an estimate and associated uncertainty interval for the infection time of each individual in the study.

The pNLME modeling approach provides several advantages. By using a population model, it is possible to estimate infection times in individuals with sparse viral load data, including those without any measurements during viral upslope. Viral dynamics are not as sensitive to multiple founder infections as evolutionary methods. And finally, mechanistic models have been used to describe viral dynamics during broadly neutralizing antibodies therapies^12^, suggesting our methodology might be applicable to infection time estimation from emerging trial data including a therapeutic prevention modality.

## Results

### A framework for estimating infection time using viral dynamics

Using experimental data and modeling, we set out to develop a framework for estimating HIV infection time from viral load data. Using observed first positive viral load, we worked backwards toward infection time and defined several precise moments during HIV primary infection for modeling (**Fig 1**). HIV infection begins with an *infecting exposure*, the target time of estimation. Starting with this exposure event, there is a brief “black-box” phase encompassing biology not captured with past viral dynamic models. For example, the virus may need to diffuse or clear mucosal and anatomical barriers before beginning viral replication as described by mechanistic models. Animal challenge studies and human cases where infecting exposure is almost certainly known suggest this period is brief, from a few hours to 1 day^9,13,14^, but given the lack of information we note it here as a fundamental uncertainty and potential bias in our estimates.

**Fig 1.**
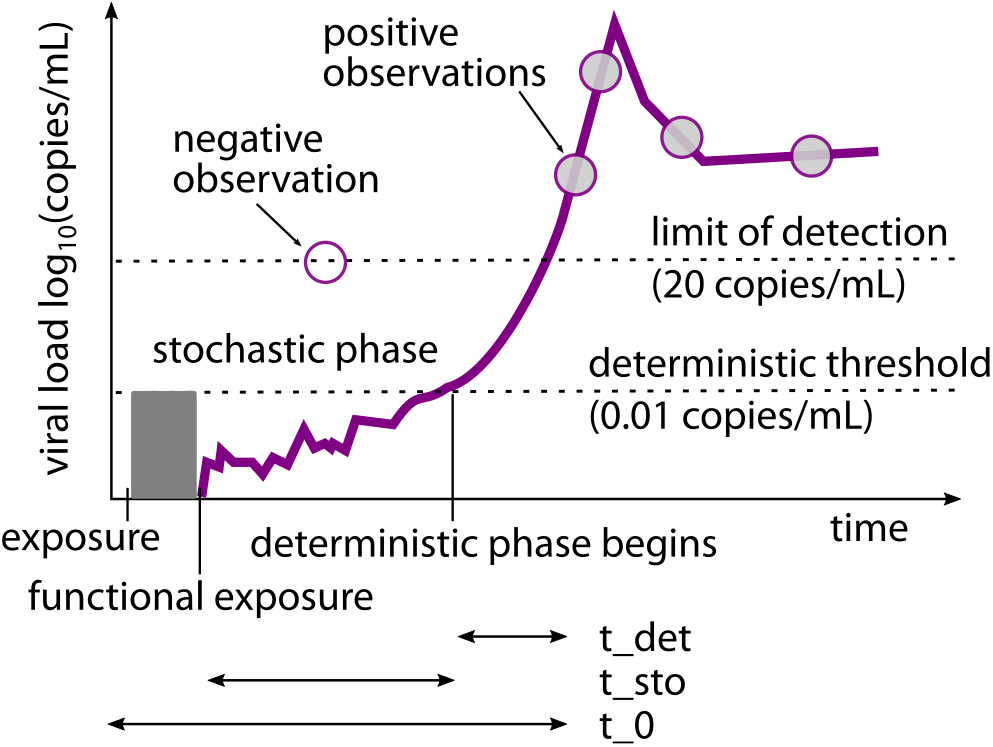
Cartoon schematic of modeling definitions. *The time between infecting exposure and first positive viral load can be described in 3 phases. First, we recognize the possibility of an unknown but likely brief “black-box” period describing localized biology that exists immediately following infecting exposure. We assume this period is short compared to the following phases. Second, a stochastic process governs early viral expansion, starting with one or a few infected cells initiating systemic infection in the new host – and concluding when viral load reaches the deterministic expansion threshold (t_sto_). Third, a deterministic model (t_det_) proceeds, describing the observed viral dynamics. By combining estimates for these phases, we finalize our estimate of t*_0_*, the time between infecting exposure and first positive viral load, sometimes referred to as the eclipse period*.

Next, we assumed that viral kinetics can be described by a dynamic model unifying observable and unobservable viral loads. At this point, we assumed bottlenecking has resulted in at most a few infected cells starting a productive infection in the human body. Viral expansion from these few cells begins a stochastic phase lasting a duration defined as the stochastic phase, *t_sto_*. The stochastic phase likely encompasses the initial viral replication at the infection foci and the transition from a localized infection to an infection that has reached germinal centers. The stochastic phase ends when the viral load crosses a *deterministic threshold*. We estimated this threshold through repeated stochastic simulations finding the minimum viral load where the 1) the slope of stochastic viral loads were nearly log-linear and 2) there was effectively no chance of stochastic burn out. We determined that a value of 0.01 copy/mL sufficiently satisfied both criteria. The time between viral load crossing the deterministic threshold and reaching the first positive viral load observation was defined as the *deterministic phase t_det_*. Finally, the time between infecting exposure and first positive viral load, comprising these three phases, we cumulatively refer to as *t*_0_. This time interval has been referred to as the *eclipse phase*^5^.

### Experimental data for model development

We used viral load observations from the RV217 study^8^ including 46 individuals out of 155 total diagnosed acute HIV-1 infections in the study. Individuals had twice-weekly HIV tests before diagnosis using the APTIMA HIV-1 RNA Qualitative Assay (Hologic)—a fingerstick device testing small blood collection (0.5 mL). Once diagnosed (2 APTIMA positive visits), quantitative PCR was used to quantitate HIV RNA twice weekly in these individuals, who did not initiate antiretroviral treatment (ART) and had ~10 study visits in the first month after diagnosis. From this cohort, we assembled viral loads from Thai and Ugandan men, women and transgender individuals. Only individuals with more than 3 detectable longitudinal viral load observations were included. We found that in very early infection, APTIMA and viral load were strongly correlated (**Fig 2A**) and APTIMA measurements could be used to impute viral load at diagnosis times for individuals without measured viral loads (**Fig 2B**). Using this relationship, we imputed viral load at APTIMA diagnosis for 28 participants, which adjusted the first positive viral load by a few days. Several individuals were not diagnosed until later acute infection, meaning that peak and upslope of viral load are not obviously detected. We do not exclude these individuals, instead relying on our population modeling approach and borrowing strength across the cohort to make estimates. These estimates are particularly useful because such data sets provide significant challenges with other modalities.

**Fig 2.**
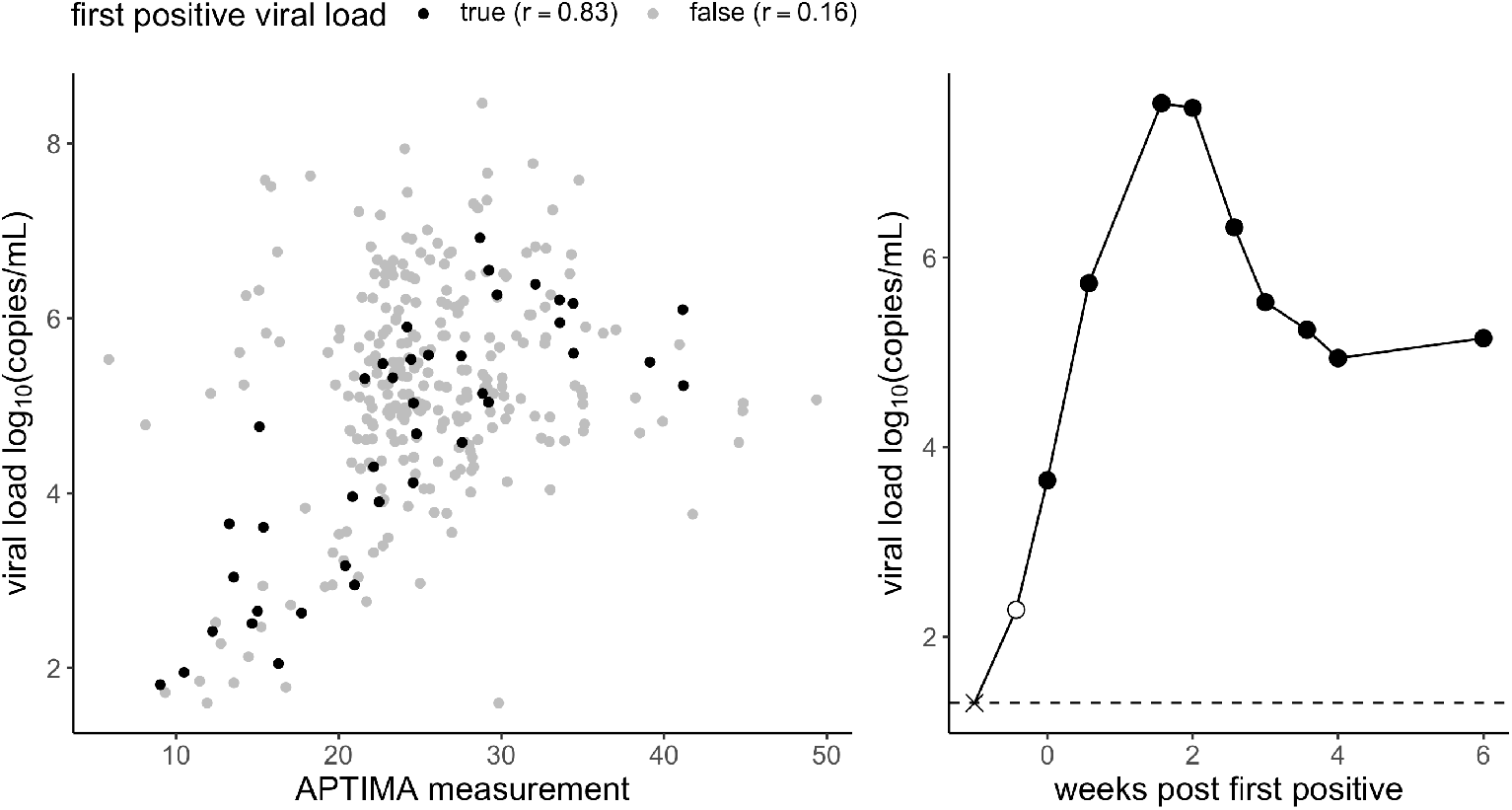
Correlation plot between APTIMA measurements and viral load. *A) A strong linear correlation (Pearson r =0.83) is found between APTIMA and log viral load at the first positive viral load (black dots). If positive samples beyond the first positive (gray dots) are included, and at higher measurements of either outcome, APTIMA is less correlated to viral load. B) We used diagnostic APTIMA measurements prior to first positive viral load to impute additional viral load values. Here the closed circles indicate observed viral load measurements, the ‘x’ the last negative measurement, and the open circle indicates an APTIMA imputed value*.

### Inference of *t_det_* from a parsimonious model to the RV217 cohort data

The first step in estimating *t_det_* was developing a model that best-described the observed data. Thus, we selected four distinct and previously applied mechanistic models of HIV primary infection and varied their population parameterizations (the number and type of parameters estimated). This resulted in a total of 30 models (see **Supplementary Table 1**). The four mechanistic models included the canonical viral dynamics model^15^, two models recently fit to SHIV/SIV viral dynamics^16,17^, and our own simplified model based upon Ref^18^. We found that the most parsimonious model to the RV217 cohort data (**Supplementary figure 1 and Table 1**) includes susceptible target cells (*S*) that are born and die naturally and virus (*V*) that infects these cells and creates productively infected cells that produce viable virus (*I*). Infected cell death rate depends on their own density powered by an exponent (h). This term semi-mechanistically encapsulates natural cytopathic cell death during viral production, as well as innate or acquired immunity against HIV infected cells that escalates as the number of infected cells increases (**Fig 3A**, see also **Methods** Eq. 1). In this way, an explicit immune effector compartment is not needed, and the model is simplified substantially.

**Fig 3.**
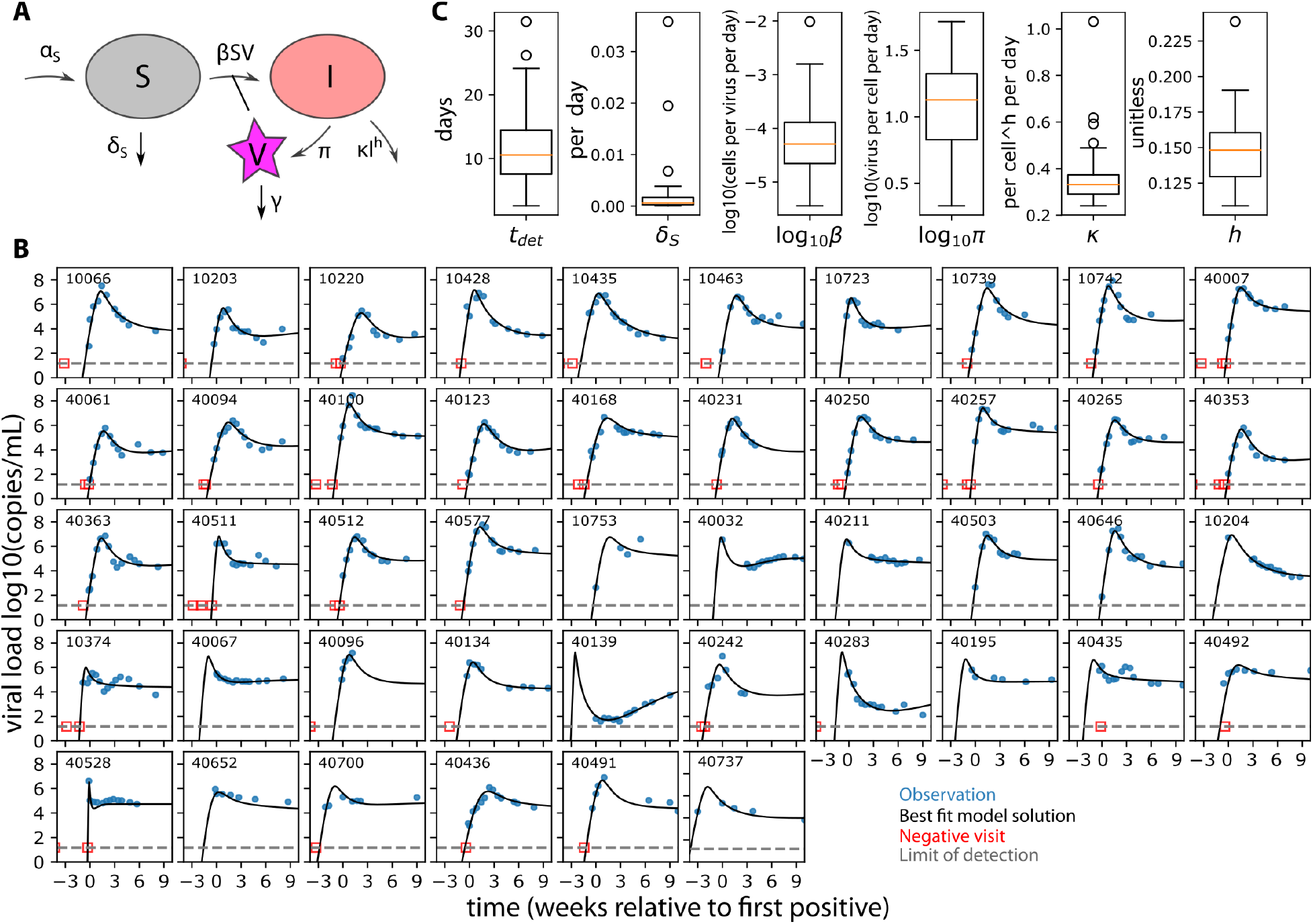
The optimal mathematical model recapitulating RV217 viral load kinetics. *A) By comparison of four structurally distinct models and many distinct statistical population models for each, we effectively tested 30 models for the data and arrived at an optimally parsimonious model, schematized here. The model is identical to the canonical viral dynamics model except infected cells have a nonlinear death rate (see Eq 1). B) This model recapitulates diverse viral load kinetics in the RV217 human study. In each panel, viral data are gray dots and best individual fit is a blue line. By borrowing strength through the population fitting approach, the model infers peak and upslope even when those data are missing (see 40139, or 40700 for example). Last negative visits are shown as viral loads at the limit of detection (20 copies/mL) and were included as censored data for fitting. Only 1 individual (last panel, 40737) had a first positive viral load that would be shifted substantially given the APTIMA imputation. C) 6 parameters were estimated including the deterministic infection time, the crucial variable for timing infection*.

The model output is congruent with previous data for other model compartments. For example, it predicts a susceptible cell drop between 40—80%^19^ (which may relate to the CD4+ T cell depletion during peak viremia^20^) and allows for the large observed inter-participant variation of viral peak (**Supplementary figure 2A**).

The best fit model for each individual is displayed in **Fig 3B**. We used population nonlinear mixed-effects (pNLME) modeling to estimate parameters, such that each individual has their own estimated parameters, but these estimates are constrained to be drawn from population distributions of each parameter; the population distribution is simultaneously estimated. All distributions of parameter estimates are shown in **Fig 3C** and values are quoted for each individual in **Supplementary Table 2**. From this model, the estimated time between deterministic threshold and first positive viral load, *t_det_*, (**Fig 1**) ranged from 2.5—32.6 days across the 46 participants with a median of 10.1 days. We also verified that models with comparable AIC (<10 difference from the best model AIC score) predict similar individual values for *t_det_*. Summary statistics of viral load (peak and set point) were not correlated with deterministic time *t_det_*; rather they were strongly correlated with estimated infectivity (*β*), viral production rate *(π)* and the nonlinear death exponent (*h*) (**Supplementary figure 2B**). The magnitude of the first positive viral load was significantly, but not strongly, correlated with *t_det_* (**Supplementary figure 3**). These results suggest other estimated parameters are mostly independent of infection timing and that the model predictions are informative beyond upslope regression-i.e. nonlinear estimation enhances our predictive power.

### Stochastic simulations until the deterministic threshold

Evidence from modeling other viruses suggests that early stochastic events are linked to later deterministic kinetics^21^. For example, for cytomegalovirus (CMV) infection, extinction probabilities, duration, and magnitude of transient stochastic infections are consistent with primary infection mathematical model parameters^22^. Therefore, based on the individual best fit parameter sets, we performed stochastic simulations to determine the time-window between the introduction of a single infected cell and the deterministic threshold *(t_sto_* in **Fig 1**). Simulations were initialized with a single infected cell per μL *1(0*) = 1 and at the viral free equilibrium between susceptible cell birth and death S(0) = *vol* × *a_S_/δ_S_*. Scaling up to realistic volumes allows for a discretized stochastic simulation; *vol* was chosen to be 5 × 10^8^ μL, or 5 L of blood (typical for adult human) at approximately 100-fold concentration based on the finding that the majority of lymphocytes reside in lymphoid tissues where infection is assumed to initiate before spilling over into blood^9,23^.

For each individual, the best fit parameters of the deterministic model were used to conduct 10 stochastic simulations via the tau-leap method^24^. Because HIV transmission is a rare per coital event^25^ and we are interested in infection time estimation, we conditioned upon successful infection^10^ by only using simulations from stochastic runs that did not go extinct. The simulations were halted when viral load crossed the deterministic threshold (0.01 copies/mL) and the time to reach that viral level *(t_sto_*) was recorded. Simulated viral loads from a single stochastic simulation of each individual are shown in **Fig 4A**. The distribution of stochastic times *(t_sto_*) is vizualized above the viral load panel, indicating a slightly asymmetric time to crossing the deterministic threshold with median ~5 days in this single stochastic simulation. There is substantial variability in the slope of these viral load trajectories based on the range of parameters inferred from the deterministic model for each individual. We also performed replicate simulations for single individuals (10 replicates for participant 10428 are shown in **Fig 4B**). In this case, viral load slopes are nearly identical by the time the deterministic threshold is crossed, but the early stochastic events introduce some variability in *t_sto_*. For this individual, the median time between infection and deterministic threshold was 5 days, with total range between 3-5 days in these 10 simulations. In summary, viral load upslope varies highly across subjects but minimally within-subjects. Variability introduced by the stochastic phase is predominantly a shift, rather than a scaling of infection time. This agrees with modeling of barcoded virus data early in infection (recently reported by Docken et al. during Dynamics & Evolution of HIV and Other Viruses 2020).

**Fig 4.**
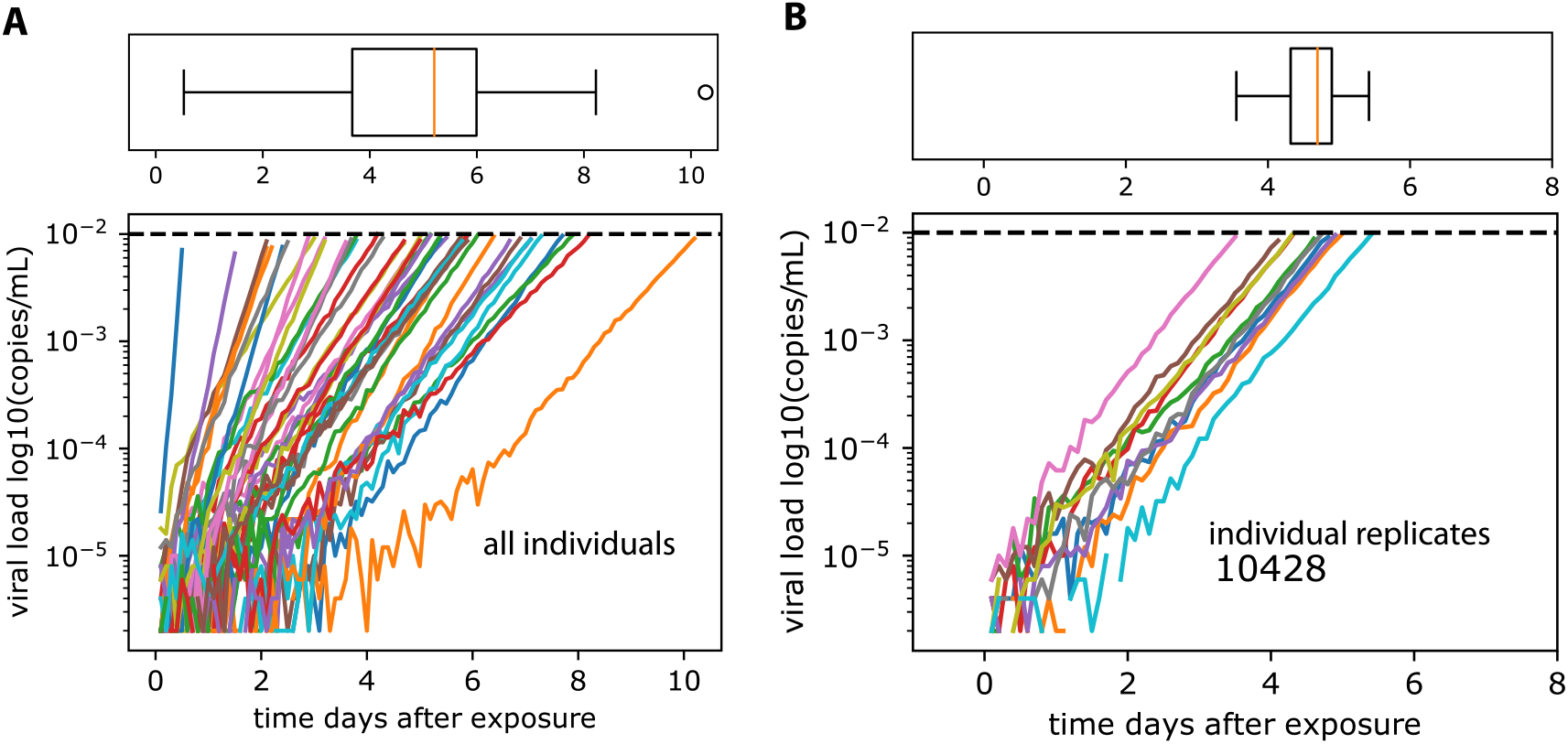
Stochastic simulations using best-fit parameter estimates from deterministic model. *Viral load kinetics until deterministic threshold (0.01 copy/mL). A) A single stochastic realization for all 46 individual parameter sets from* ***Fig 3*** *with associated boxplot of the distribution of times (of these 46 simulations) to reach the deterministic threshold. The slopes are different across individuals owing to the different parameter estimates from the deterministic models. B) 10 replicate stochastic realizations for a single individual until deterministic threshold with associated boxplot of the distribution of times (of these 10 simulations) to reach the deterministic threshold. Here slopes are nearly identical, but due to the stochasticity of the simulation, the time to reach the deterministic threshold varies between 3-5 days. Note discontinuities in lines are artifacts of downsampling for file size considerations*.

An important parameter for these simulations is the initial number of infected cells. We show estimates of *t_sto_* are inversely correlated with *1(0)*. For example, as *1(0)* was increased from 1, 10, 100, to 1000, the median estimate of *t_sto_* across individuals decreased 5-1 days (**Supplementary figure 4**). As a result, this difficult-to-measure biological parameter only adjusts estimates by a few days.

### Combining the stochastic and deterministic phases to estimate infection time

Next, we integrated the stochastic and deterministic timing estimates to complete the estimation of *t*_0_, the time between infecting exposure and first positive viral load, or the eclipse phase (**Fig 1**). Here as an example, we present this procedure for individual 10066 (**Fig 5**). First, we used the best fit parameters and performed 100 replicate stochastic simulations to estimate a distribution of *t_sto_*; the mean was approximately 6 days, and the distribution was skewed, with 95% uncertainty interval ranging between 4-9 days. Second, we drew values of *t_det_* from a constructed conditional distribution using Markov-Chain Monte-Carlo given the population and random effect estimates of *t_det_* (mean 10, 95% uncertainty interval between 7-13 days). The infection time, *t*_0_, was then calculated by drawing and summing *t_sto_* and *t_det_* from their respective distributions. This was repeated 10000 times to generate an average *t*_0_ with associated 95% uncertainty interval (see estimates of *t_sto_, t_det_*, and *t*_0_ for this individual in **Fig 5**). We estimated that this individual’s infection occurred 16 days prior to first positive viral load with 95% uncertainty interval ranging between 12 and 20 days. This procedure was performed for all individuals.

**Fig 5.**
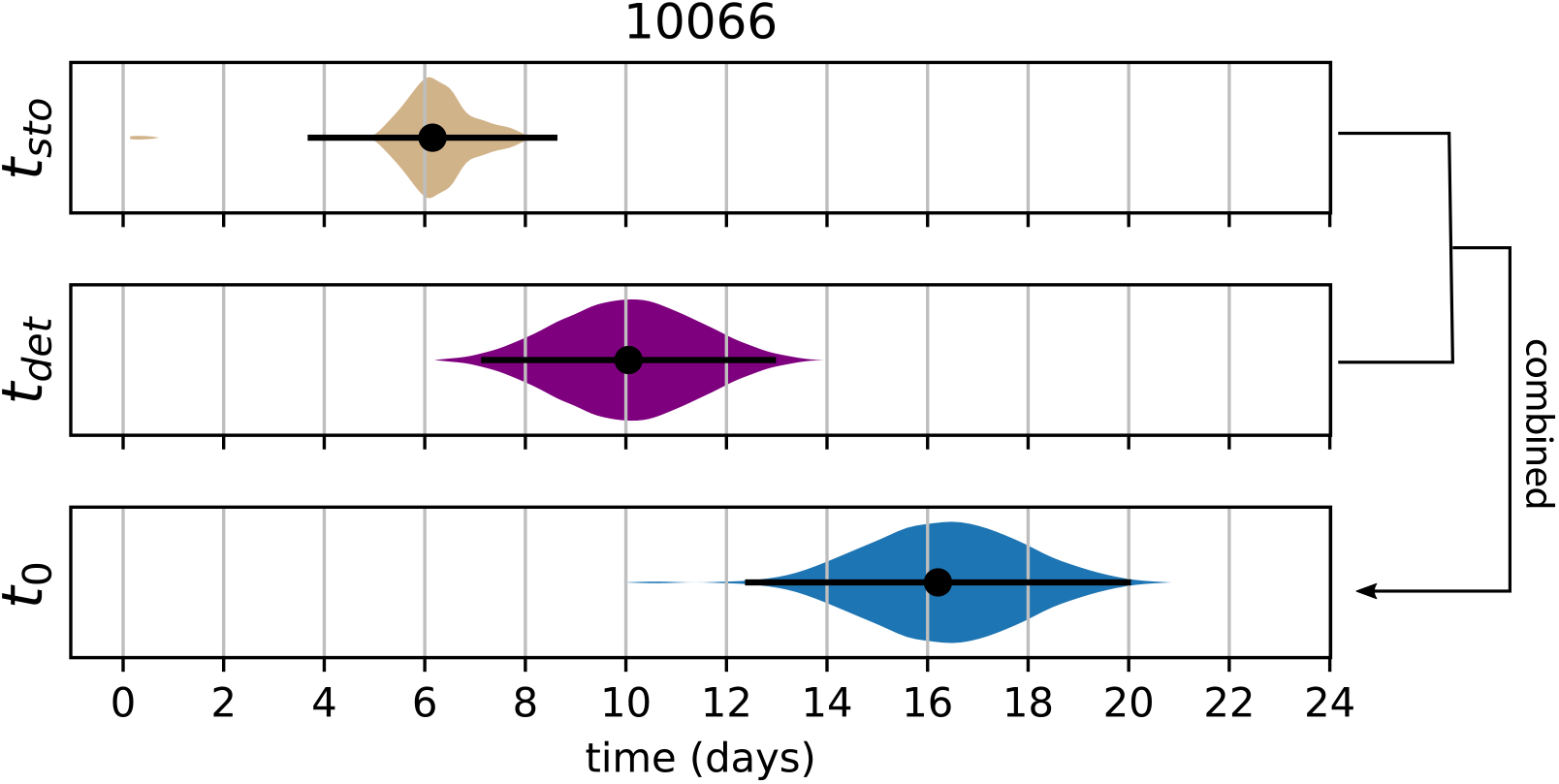
Individual estimate example. *Bootstrap combination of the deterministic and stochastic estimates provides an estimate for an individual’s (10066) time of infection. The stochastic time interval (t_sto_) between 1 infected cell and the deterministic threshold (0.01 copies/mL) was determined by 100 replicate stochastic simulation for that individual. Here the mean estimate of t_sto_ was 6 days (dot) with 95% uncertainty interval (lines) ranging between 4-9 days. The entire probability distribution is shown to illustrate skew. The time interval between the deterministic threshold and the first positive viral load was determined by the best estimate of t_det_ using population nonlinear mixed effects modeling (see* ***Fig 3****). Here the mean (dot, ~10 days) and 95% uncertainty interval (lines, ranging between 7-13 days) from the MCMC estimate are shown with the derived distribution. Finally, the time between infection and first positive viral load (t*_0_*) is calculated by 10000 random combinations of t_det_ and t_sto_ drawn from these distributions. Our best estimate suggests this individual was infected 16 days prior to first positive with 95% uncertainty interval ranging from 12-20 days*.

### Direct comparison to previously applied infection timing estimation tools

Rolland et al. used several viral load and phylogenetic inference techniques to estimate infection times using the RV217 data^3^. These methods are the maximum slope of any two points on the upslope (max_slope), the best log-linear regression slope (linear_model), self-reported entries from trial participants (self_report), Bayesian phylogenetic inference of median time to most-recent common ancestor (BEAST)^26^, and Poisson fitter^27^ diversity estimator based on envelope sequences sampled at three time points in the first six months of infection. We compared our viral dynamics population non-linear mixed effects (pNLME) modeling approach estimations directly against all methods in **Fig 6**. In general, our deterministic estimates were in the same relative range of the other estimates. However, concordance correlation coefficients (CCC), which score how close data lie to the line y=x, are in general fairly weak (CCC<0.4) between pNLME and other methods (**Fig 6A**). This is driven by the fact that the complete estimator finds infection time earlier than most other methods, perhaps due to the additional stochastic phase. Adjusting the initial number of infected cells from 1 to 1000 (see **Supplementary figure 4**), or removing the stochastic phase decreases the average eclipse time, closer to previous estimates. However, we show correlation between pNLME with and without the stochastic component (final panel in **Fig 6A**) to illustrate this relationship is not necessarily linear.

**Fig 6.**
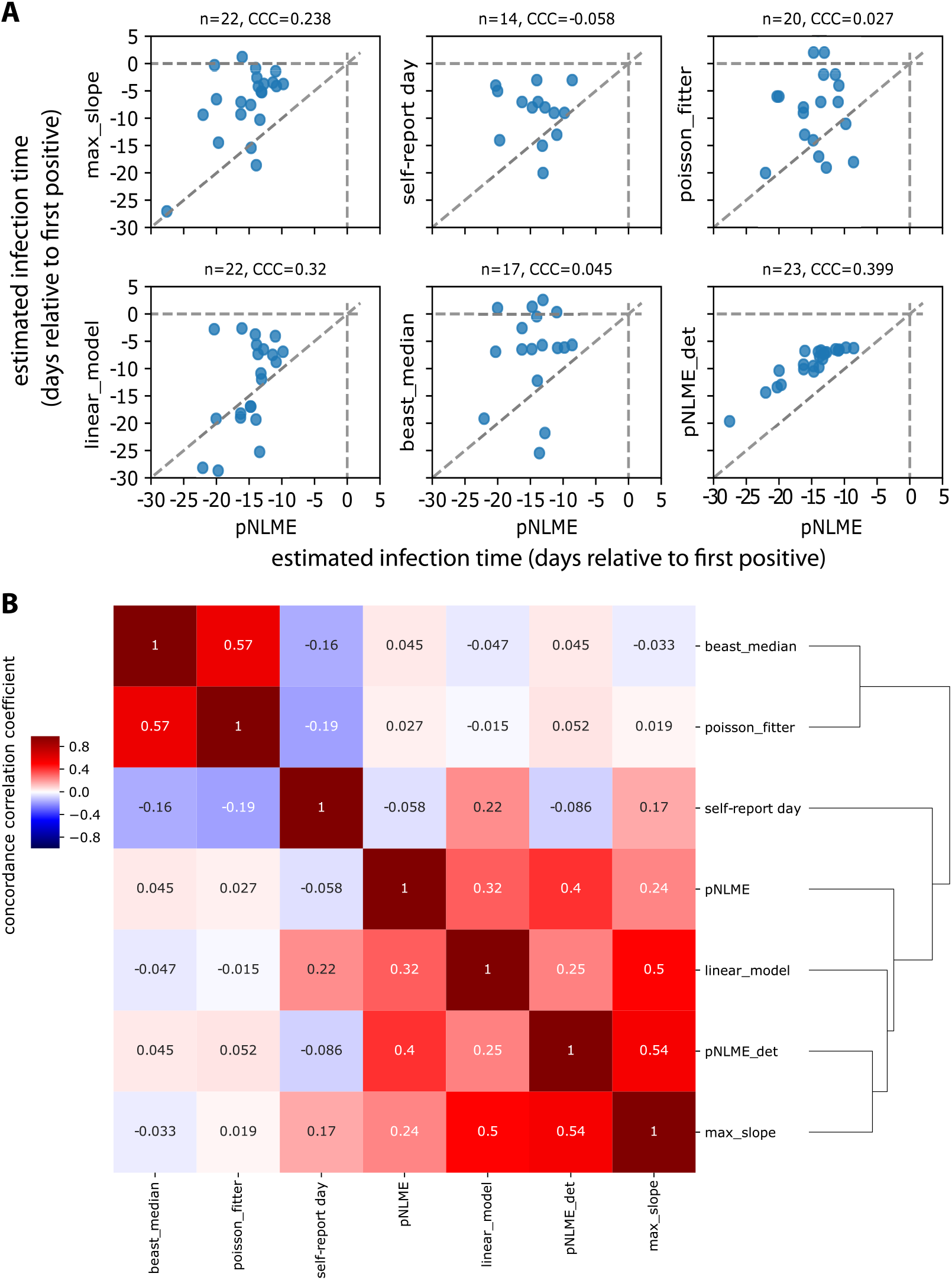
Comparisons of the population non-linear mixed model estimation (pNLME) approach for infection timing versus 5 other methods. *Methods include the maximum slope of any two points on the upslope, the best log-linear regression slope (linear_model), self-reported entries from trial participants, Bayesian phylogenetic inference of median time to most-recent common ancestor (BEAST), Poisson fitter diversity-based estimator, and our own approach using only the deterministic component. A) Best estimate of each available individual from each method expressed as predicted infection time relative to first positive (all estimates are tabulated in* ***Supplementary Table 2****.) An estimate above 0 (see dashed lines), indicates unrealistic estimates of infection after first positive. While our method can be used on all 46 individuals, other approaches are constrained by features of the data (e.g. detection of upslope, sequencing characteristics). Thus, comparison size n is denoted above each panel. Concordance correlation coefficients show individual agreement is generally weak: CCC = 1 when all data lie on the line y = x (shown as a dashed line). B) Hierarchical clustering using concordance correlation coefficients indicate which methods give most similar estimates. Sequence based methods and viral dynamic methods fall into 2 main clusters, with self-report falling in the middle of these. pNLME agrees more strongly with other viral dynamic methods*.

We also compared all previous point estimates to one another (**Fig 6B**). No approaches were very strongly correlated by CCC. Hierarchical clustering of previous methods shows there are two distinct groups that include genetic estimators (BEAST, PFitter) and viral dynamic estimators (max-slope, linear_model). Self-report diary entries and our method (pNLME) fall roughly in between.

### Wide applicability of pNLME to sparse data

The pNLME approach is widely applicable to data that challenge other methods. It can provide estimates for individuals for whom viral upslope is completely undetected. It also does not produce large outliers and never estimated the time of infection to be after first positive, as Max-slope, BEAST, and PFitter do in a few cases. pNLME also does not appear to be sensitive to multiple founder infections (which are particularly difficult for genetic estimators). For example, Rolland et al. identified some individuals in this cohort infected with multiple founder viruses based on the sequence analysis; for these infections, estimates of time to most recent common ancestor often gave estimates preceding the date of last negative test by many months (reflecting divergence in the transmitting partner rather than divergence after transmission)^3^. Infection with multiple founders has been associated with higher set-point viral loads^28^. Therefore, we tested to see if our model parameter estimates were different in single versus multi-founder infections (as differentiated by Rolland et al., see **Supplementary figure 5**). We observed no obvious patterns distinguishing single and multi-founder participants and found no significant differences among our parameters (Mann-Whitney p>0.1) but note the limited sample size with these data (n=9 multiple founders in this set). While beyond the scope of this paper, applying multi-founder status as a pNLME model covariate might admit more power given the small sample size. Importantly, the estimate of the time of theoretically crossing the detection limit was not affected by the distinction of multiple founders, meaning our estimates are robust to this challenge for phylogenetic inference.

### Proof of concept study on synthetic data with realistic study protocols

To assess the accuracy of pNLME estimates, we performed a simulation study using several different sampling schemes. We simulated viral loads from 20 randomly chosen RV217 participants and sampled these viral loads with 5 different theoretical protocols. The first we refer to as “gold” meaning daily sampling before and after first positive. We refer to tight as weekly sampling visits, and sparse as monthly sampling visits (every 4 weeks). Infection was assumed to occur uniformly between study visits. If viral load was above 20 copies/mL at a visit that sample was called first positive (or diagnosis, dx), and measurements occurred subsequently. In **Fig 7A** we illustrate an example infection (red x), viral load (blue line) and observations (orange circles) for each protocol: “tight pre / tight post”, “tight pre / sparse post”, “sparse pre / tight post”, “sparse pre / sparse post”, and “gold”. We took these synthetic data observations and estimated *t*_0_ with pNLME. In this step, we used the RV217-trained model, meaning that we fixed the population distributions (as we would with new test data), and arrived at a new conditional distribution of individual parameters for each synthetic data set. We applied those parameters to the stochastic modeling step, completing the estimate of *t*_0_ on each synthetic data set.

**Fig 7.**
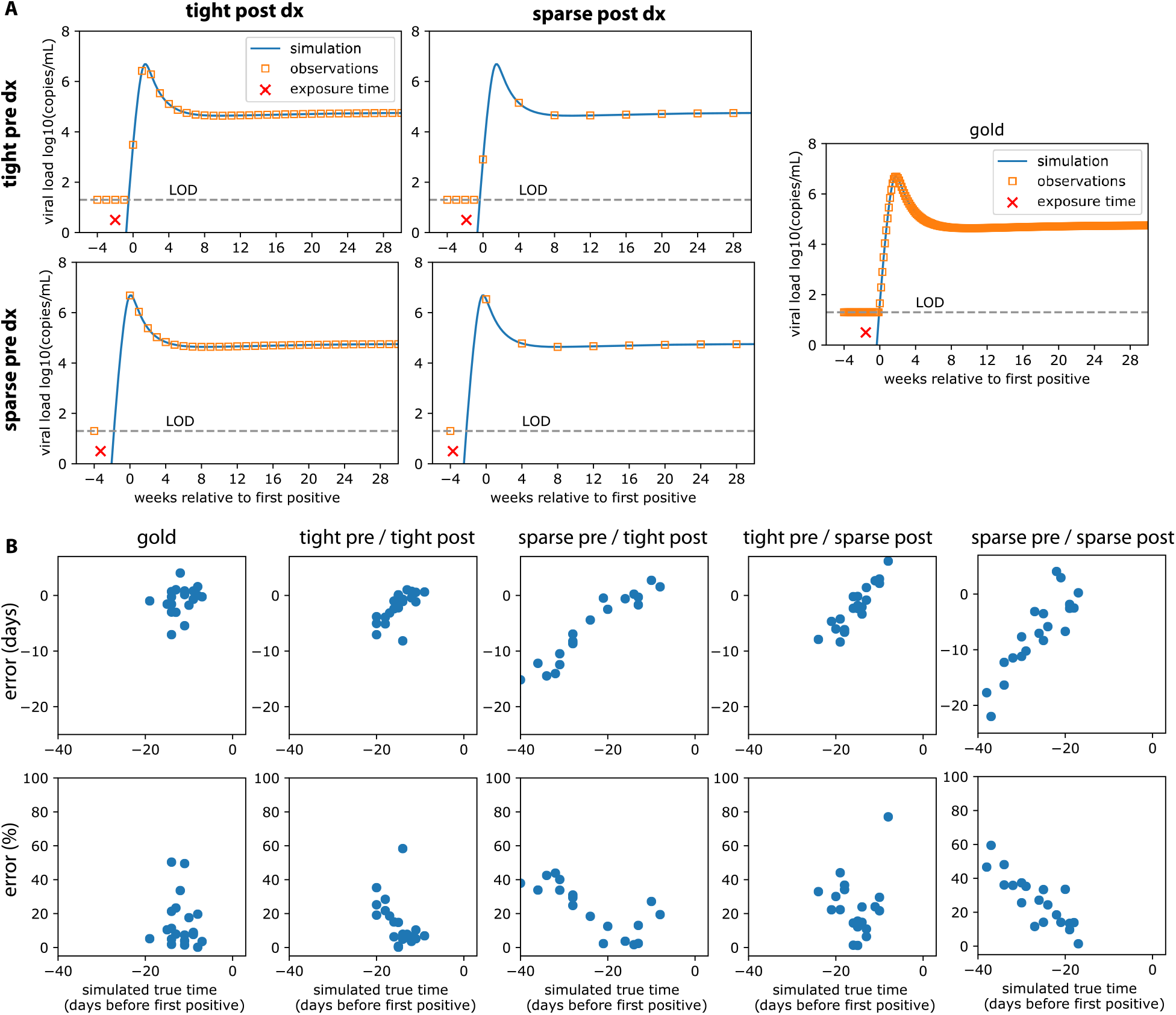
Accuracy of timing tested on simulated viral load data with sparse and tight study sampling. *We simulated viral loads using individual parameter sets and sampled according to 4 different theoretical study protocols. We refer to tight as weekly, and sparse as monthly (every 4 weeks). We simulated data assuming infection occurred uniformly throughout observations periods. If viral load was above the study assay detection limit at an observation (20 copies/mL), that was called first positive (or diagnosis, dx), and measurements occurred subsequently. A) 4 examples of each study protocol: combinations of tight and sparse, pre and post diagnosis (dx). True infection is denoted with the red x, simulated viral load with the blue line, and observations given the protocol with the orange squares. B) 20 individuals were simulated with each protocol, and pNLME inference was performed on those data. The accuracy of the estimated t*_0_ *compared to the true infection time is shown as error in days (difference between pNLME estimated and true time) and percent error (error relative to true time x 100%) for each sampling protocol. Low error therefore indicates estimates that agree, and percent error illustrates how estimates get more biased (relatively to other estimates) as the time between detection and true infection time increases*.

**Fig 7B** shows the absolute error (days difference between truth from the synthetic data and inferred *t*_0_ from pNLME applied to those data) and the % error: (true-inferred)/true x 100%. Gold standard and tight/tight predictably admitted the lowest errors. Very few individuals were overestimated, meaning that when inference was incorrect, their infection time was usually closer to first positive than inferred. The exception to this occurred for some individuals with sparse post sampling.

All schemes other than gold had an obvious bias. Absolute and percent error was higher in individuals for whom true infection time was earlier. This means that uncertainty rises with estimation time farther from first positive. Put another way, our confidence decreases as the estimator projects farther into the past- an intuitively satisfying, albeit challenging finding. That this effect was fairly linear hints that it might be corrected. However, this may be an artifact of our synthetic data exercise, so we opted not to follow through with any correction. Rather, we focus on individuals who appear to have been infected within 20 days since first positive. For all sampling schemes, error on these estimates has a median of +/-10 days. A corollary of this finding is that sparse sampling after diagnosis was less detrimental than sparse sampling before diagnosis, because of the growing uncertainty with time and the likelihood of missing upslope, peak, and downslope.

This exercise illustrates one of the most practical applications for this method: estimating infection time in clinical trials of HIV pre-exposure prophylaxis agents. The “sparse/sparse” case represents a protocol comparable to that of the AMP (antibody mediated prevention) studies. In that study, visits occur every 4 weeks, and after first positive (week 0) visits occur at week 2, 4, 8, 12, and 24 weeks^12,29,30^. Thus, given the data generation distribution produced under our modeling assumptions, and the additional assumption that HIV dynamics in participants in the AMP study are comparable to participants in RV217, we expect our approach would provide reasonably accurate estimates for individuals who appear to have been infected within 20 days of first positive visit (95% uncertainty interval ~5 days), and less confident estimates for others. A secondary result of our modeling is that more sensitive detection could be crucial to avoiding the ultimately challenging case of individuals infected >4 weeks before first positive visit. APTIMA testing and viral load prediction as in **Fig 2** would help this substantially.

## Discussion

Estimating infection time is especially critical in HIV prevention trials. If drug levels at the precise time of infection can also be estimated then required drug levels for protection may be identified. Here, we have demonstrated estimation of HIV infection time using non-linear viral dynamics. Specifically, we assess the viral load trajectory, an established endpoint for HIV trials. We developed a two-step procedure: 1) using population non-linear mixed effects (pNLME) modeling to estimate parameters for individuals who were infected with HIV and then 2) using these same parameters to repeatedly simulate stochastic infections. In step one, we estimate the time between first detected positive viral load and the viral load reaching some level theoretically considered deterministic. In step two, we quantify the time of stochastic viral growth until the deterministic threshold. Combining steps 1 and 2 completes the estimate of the time between exposure/acquisition and viral load detectability, sometimes called the “eclipse time”.

We applied our method to data from the RV217 observational cohort, an acute HIV infection study with highly granular measures of viral load early during infection. For the first step we performed extensive model selection and found a mechanistic model that recapitulates viral loads in the RV217 trial from first positive until viral set point. A trained pNLME model using data from multiple individuals that includes observations during several stages of viral infection (e.g. expansion, peak and set point) allows confidence in parameter estimation from individuals who may not have data from all stages.

We compared our technique to other techniques that were applied to the same data set. We find that the individual level estimates are not concordant. On the population level, average values of our deterministic model agree with average values from other approaches. The additional stochastic phase in our model drives our estimates slightly farther from time of first positive, extending the range of the eclipse time. Concordance of our model is strongest (CCC=0.2-0.4) with other approaches that use viral load dynamics. Sequence-based approaches are the least concordant, and self-report diaries are somewhat middling. We note that the study group remains was too small to evaluate certain variables that differed across individuals, such as viral subtype, sex, age or ethnicity. In cases where viral dynamics and sequencing data exist, it may be optimal to try all approaches and developing uncertainty intervals extending across all methods. A future solution would be to include evolution into our mechanistic model and fit to both types of data.

Compared to other methods, our approach has several advantages. It allows estimation of infection time in individuals without well resolved viral upslope or even viral peaks. Specifically, without incorporating the population data, it is not possible to estimate infection time when viral upslope is missed. It is also relatively insensitive to founder multiplicity, a challenge for phylogenetic methods that sometimes results in unrealistic infection time estimates after the first positive viral load. The RV217 study is unlikely to be repeated. Thus, our model can be considered a trained model for future trials. Moreover, our synthetic data sampling study illustrates what such trials might look like.

While the true time of infection cannot be known other than in challenge experiments. We verified that our RV217-trained pNLME model works on simulated data (from the same mechanistic model). Even given a sparse sampling scheme (0,2,4,8,12,24 weeks after first positive) as in the antibody mediated prevention (AMP) studies, the approach generally works well for individuals for whom infection estimates are less than 20 days before first positive. This means that protocols sampling with ~2-3 week intervals typically have <20% error, or at worst 7 days off. However, our uncertainty grows for infections occurring further before first positive. This reflects the challenge of estimating data such as sparse/sparse in **Fig 7A**. Collecting only setpoint, or partial downslope means the estimate relies heavily on the population model, and with the heterogeneity of individuals, can be relatively error prone. Tighter sampling after first positive does not drastically improve accuracy. Indeed, for trial design, accuracy is better-enhanced by tighter sampling prior to diagnosis.

The largest challenge for the approach occurs when individuals are infected at a study visit (these occur every 4 weeks) but are not diagnosed because their viral loads are below detection, meaning that the first positive viral load will be not detected until >4 weeks after infection. This challenge is not unique to our approach, and we stress diagnostic-focused assays such as APTIMA to make diagnoses as soon as possible in situations where infection timing is crucial. Thus, we have shown that 1) it is possible to leverage and impute viral loads based on the finding that early APTIMA measurements are correlated to qPCR measurements, and 2) that borrowing strength using population modeling may be the best option to overcome sparse sampling.

There are several limitations to our study. It remains unknown, and will be extremely hard to test, whether early HIV dynamics can be described by the same mechanistic model as deterministic viral dynamics. However, in CMV transmission the probability of infection has been related to post-infection viral kinetics, suggesting stochastic behaviors may be linked to subsequent deterministic kinetics^22^. We speculate an early lag period in HIV infection that could be described by localized exposure and viral escape from anatomical barriers before initiating systemic infection. The duration of this period is unknown and we effectively assumed that it is negligible compared to the stochastic and deterministic phases, and compared to our window of estimation (i.e., less than a day). To account for this lag period, a further non-mechanistic window might also be added to reconcile wider estimates of eclipse time found in some studies^6^. Of note, it is not clear if any timing method can directly account for this period; for example, the founder sequence may describe the virion that escapes the early barriers in our schematic.

Our choice of the initial simulation conditions *1(0)* inversely correlates with time of infection. That is, if we assume viral infection begins at a lower level, our estimates are further back in time. However, in what we consider to be a plausible range of initial conditions (ranging from 1 to 1000 infected cells initiating infection), the estimation varies by ~5 days. Interestingly, Rolland et al. found that if using a log-linear upslope modeling approach, a viral load of 1 copy/mL gave the best estimates in non-human primate infection where the date of infection was known perfectly^3^. One might therefore choose this value for the deterministic threshold, but the translation of this estimate is limited by the NHP experimental model, challenge virus species, and differences from viral load exposures in human transmission.

This approach should be generally applicable to other viruses. For example in Hepatitis C mechanistic models have been developed and some prior parameter estimates have been recorded^31^. Recently a similar method was applied to estimate the time of SARS-COV-2 infection^32^.

In future work, we plan to explore modifications due to preventative interventions, such that timing estimation, and therefore drug efficacy can be better estimated in upcoming clinical trials using broadly neutralizing antibodies.

## Data Availability

All data and computational code are available from the authors and will be posted to github after acceptance.

## Acknowledgements

DBR thanks Paul Edlefsen for motivating this work, as well as Raabya Rossenkhan, Philip Labuschange, Dobromir Dimitrov, James Moore, Holly Janes, and Yunda Huang for help and valuable conversations. DBR is supported by the Washington Research Foundation and an NIH K25.

## Methods

### Most parsimonious mathematical model

The set of ordinary differential equations for the model that is selected for this approach can be written as

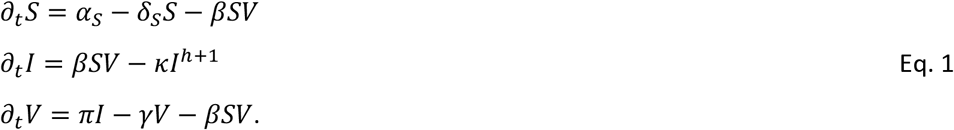

The selected model contains 8 free parameters *θ = (α_S_, δ_S_, β, k, h, π, γ,t_det_)*. The model we ultimately select is a slightly modified basic viral dynamics model that incorporates a nonlinear death term. The model tracks the concentration [cells mL^-1^] of HIV-susceptible cells *S*, infected cells *I*, and plasma viral load *V* [viral RNA copies mL^-1^]. The deterministic system is expressed (using the partial *∂_t_* to denote derivative in time) with *α_S_* [cells μL^-1^ day^-1^] the constant growth rate of susceptible cells, *δ_s_* [day-1] the death rate of susceptible cells, and *β* [μL virus^-1^ day^-1^] a mass-action viral infectivity. The viral production rate is *π* [virions cells^-1^ day^-1^], and *γ* [day^-1^] is the clearance rate of virus. The death and killing of infected cells is governed by the rate of *κ* [cells^-h^ day^-1^], with the exponential factor *h* adjusting the nonlinear density dependent death rate. This approach coarsely approximates adaptive immunity such that higher numbers of infected cells engenders faster killing.

### Population nonlinear mixed effects (pNLME) approach

We modeled the plasma viral load using a nonlinear mixed-effects approach (pNLME). In this approach an observed plasma viral load for individual *i* at time *j* is modeled as log_10_ *y_ij_ = f_V_(t_ij_, θ_i_) + ϵ_V_*. Here, *f_V_* is the solution of the nonlinear mechanistic model for the variable describing the virus *(V)* given the individual parameter vector *θ_i_* and 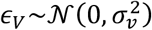 is the measurement error for the logged viral load. We assumed that the individual-specific parameter *θ_i_* is drawn from a probability distribution with median or fixed effects *θ^pop^* and random effects 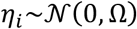, being Ω the variance-covariance matrix. Except otherwise specified we modeled parameters *β^j^* and *π^j^* as 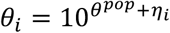 and remaining parameters as 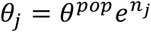.

### Model fitting

We explored four different mechanistic models with different statistical complexities, for a total of 30 models (See **Supplementary Table 1** for details). For each model we obtained the Maximum Likelihood Estimation (MLE) of the measurement error standard deviation *σ_v_*, the fixed effects vector *θ^pop^* and the elements of matrix Ω using the Stochastic Approximation of the Expectation Maximization (SAEM) algorithm embedded in the Monolix software (www.lixoft.eu). We run the SAEM algorithm 15 times (assessments) for each model using randomly selected initial guesses for the parameters to estimate. For all model fits we assumed *t_ij_* = 0 as the time of first positive viral load. However, we defined the initial value as the time *-t_det_* when *V(−t_det_) =* 0.01 copies/mL. We fixed other initial values as 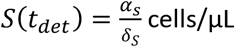 and 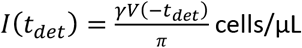 cells/μL. Per Ref ^31^, we fixed parameter *γ* = 23 day^-1^. We estimated the remaining parameters of the mechanistic model including *t_det_*. Individual parameters were selected using the mode of the conditional distribution 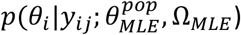 constructed by the MCMC algorithm in the Monolix software. The conditional distribution of *t_det_* for each individual is used to compute the time of infection *t*_0_.

### Model selection

To determine the most parsimonious model we calculated the log-likelihood (log *L)* for all 15 assessments for each one of the 30 models. We then computed the Akaike Information Criteria for the model with highest likelihood among the 15 assessments 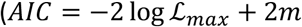, where *m* Is the number of parameters estimated). We assumed a model has similar support from the data if the difference between the AIC for its best assessment and the best one for the model with lowest AIC is less than two^33^.

### Stochastic simulation scheme

We adapted the ordinary differential equation system Eq 1 to simulate stochastically^12^. Our implementation in Python, which employs the T-leap approach^24^, is publicly available. A time interval Δ*t* = 0.0001 days is chosen for step size, in which a Poisson number of each transition type occurs. Initial conditions are changed to discrete values by multiplying by a volume. We choose this volume to be 10^8^ μL based on the observation that there is approximately 1-10 L of blood in an adult human and that there are approximately 10-100 times more T cells in lymph tissue than blood. A single infected cell is assumed (other than in sensitivity analyses in **Supplementary figure 4**).

### APTIMA analysis

APTIMA was the primary diagnostic assay in the RV217 study, of which 43 participants in our analysis were diagnosed by a positive APTIMA quantitative measurement. The remaining participants were diagnosed via a qualitative APTIMA response or directly with viral load. Among the 43 participants, only 6 had concurrent measurements of viral load for analysis. Comparing concurrent measurements APTIMA and viral load we found 1) substantial Pearson correlation between APTIMA and viral load at first positive viral load; and 2) diminished correlation later in the study at higher values of both measurements (**Figure 2A**).

Given the high correlation between the two measurements, we sought to investigate whether we could use APTIMA measurements at diagnosis to predict the unmeasured viral load for our model. This was accomplished using linear regression models predicting log first positive viral load with concurrent APTIMA as the predictor, evaluating both untransformed and log-transformed inputs (**Supplementary figure 6A**). One participant had an APTIMA measurement of 3, an outlier more than 2-fold lower than the next lowest value, and were removed from the model. To determine the appropriate upper range for APTIMA input for prediction, linear regression models were fit applying different upper bounds. Model performance was evaluated using residual mean square error (RMSE) predicting log viral load (**Supplementary figure 6B**). We found the best model used raw APTIMA measurements as the input with an upper bound of 34 (**Supplementary figure 6B&C**). This model was applied to the data to impute first positives for participants where APTIMA was measured for diagnosis without viral load (**Supplementary figure 6D**).

## Supplementary figures and tables

**Supplementary figure 1.**
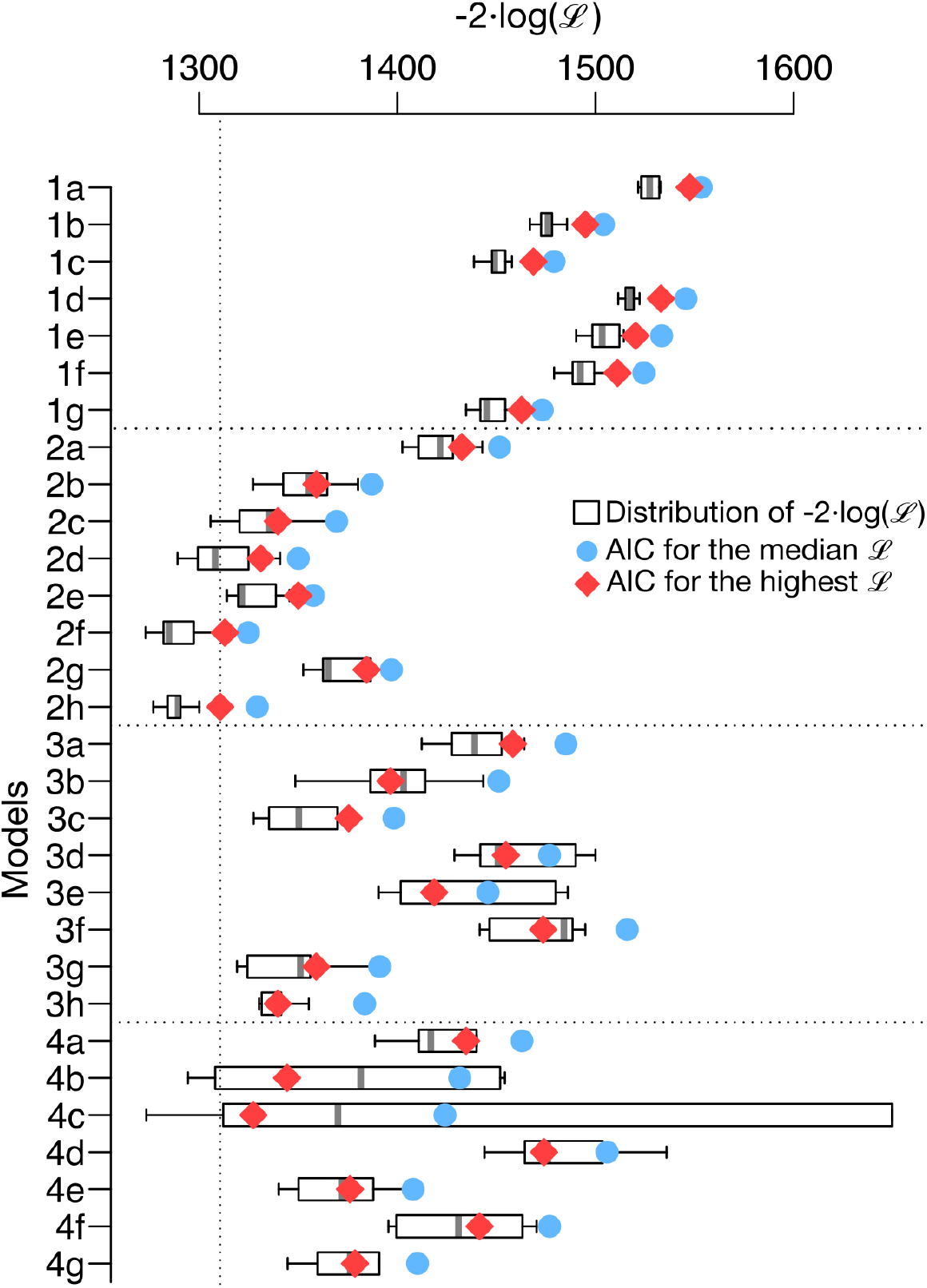
Model selection details. *All 30 models tested compared by log likelihood (-2LL) and Akaike Information Criterion (AIC). Boxplots give -2LL of 15 assessments for each model, where each assessment begins at a different parameter set and proceeds with the stochastic SAEM algorithm. Red diamonds give AIC of the median -2LL across assessments. Blue circles give the AIC of the mean assessment. The best model is 2h. Several different correlation models were attempted for each model, ultimately leading to the most identifiable model combinations*.

**Supplementary figure 2.**
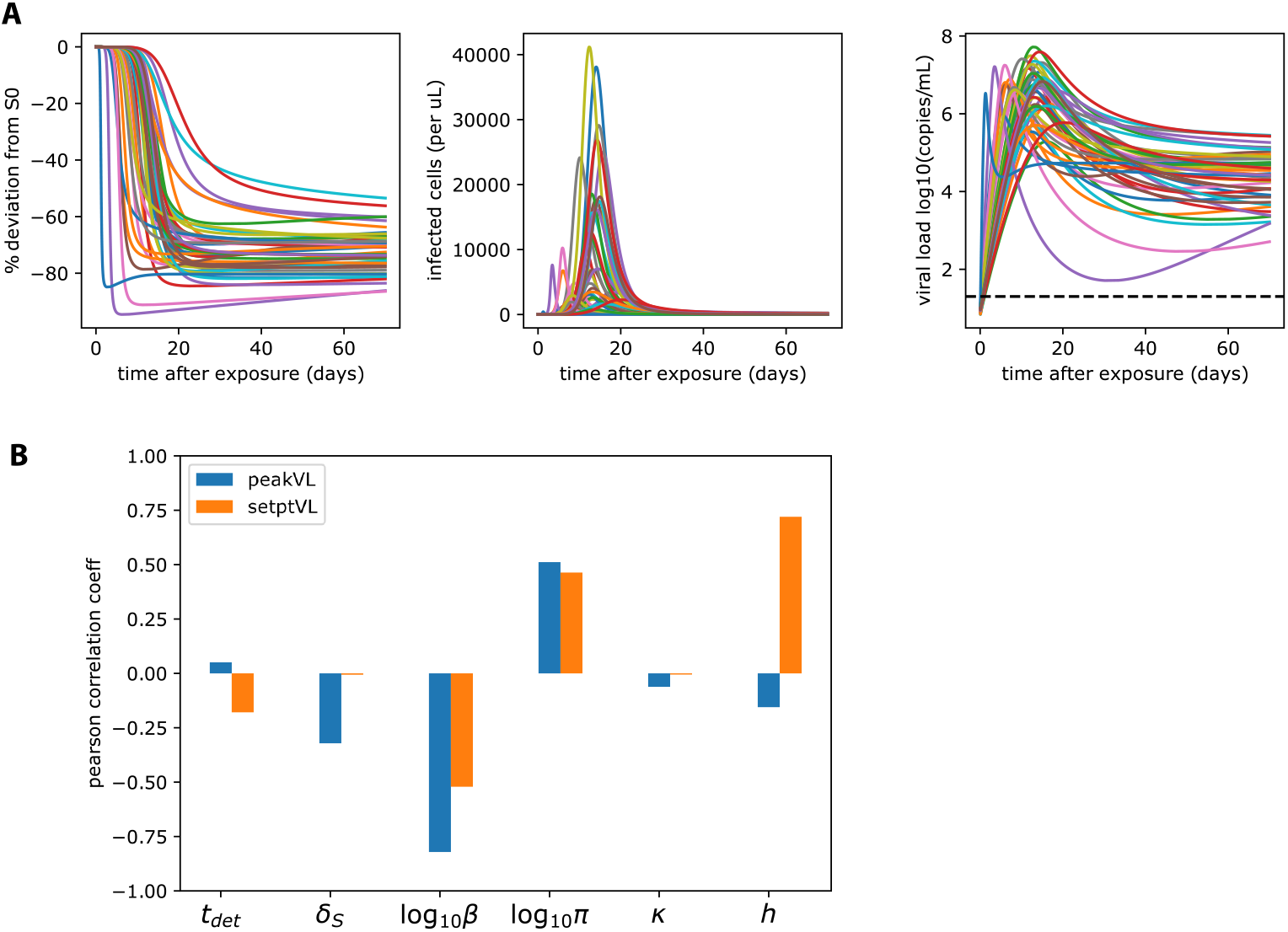
Model sensitivity analysis. *All best fit parameter sets simulated together. A) % deviation from the initial number of susceptible cells can go up to −100% percent, indicating massive destruction of cells in acute HIV infection. The total number of infected cells at that point can rise to ~1000 cells per μL. Viral loads can have peaks ranging from 10^6^-10^8^ copies/mL, with setpoints varying substantially between 10^2^-10^6^ copies/mL. B) Pearson correlation between parameters and viral kinetic phenotypes*.

**Supplementary figure 3.**
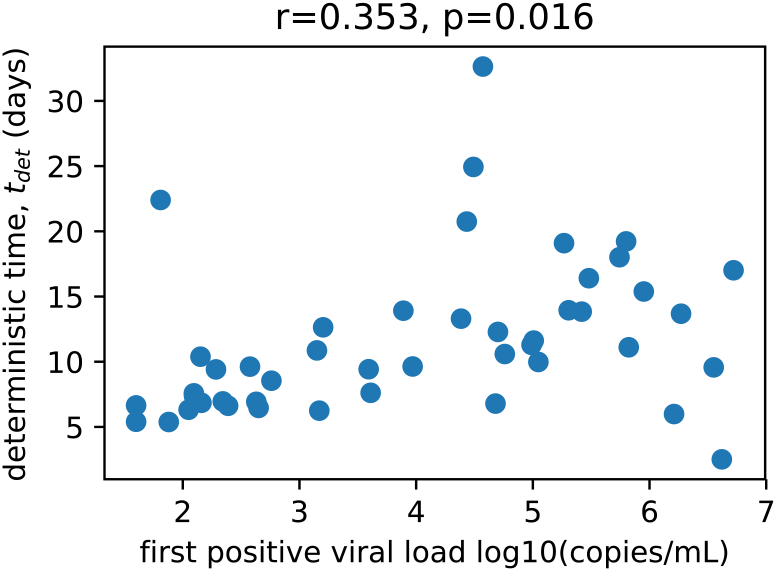
*Correlation between observed first positive viral load and the inferred deterministic time*.K0)=1

**Supplementary figure 4.**
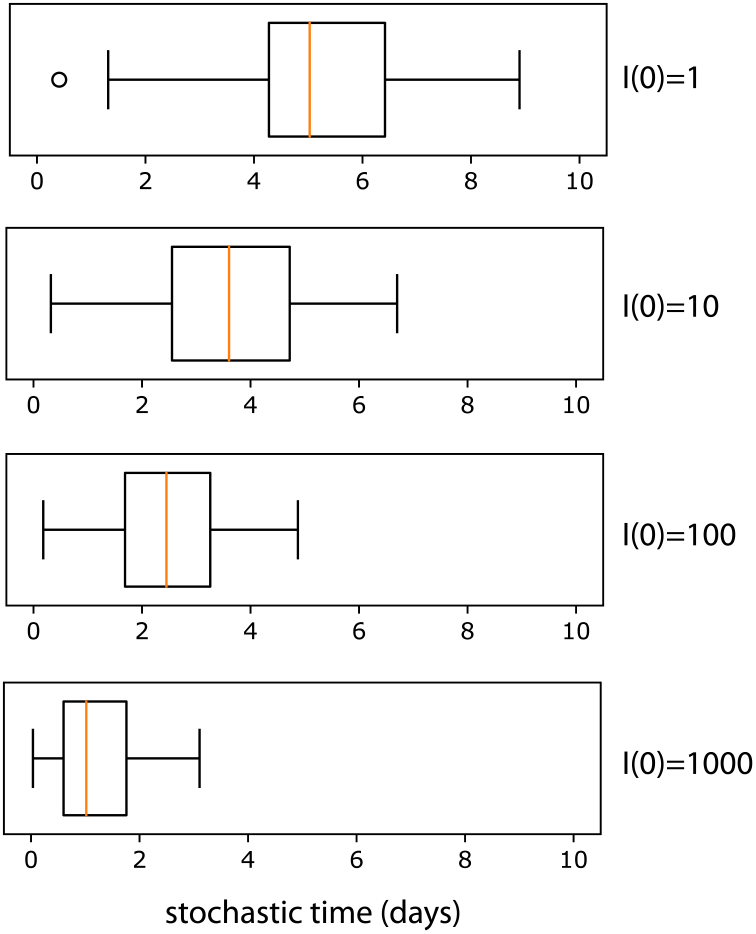
Relationship between initial number of infected cells and stochastic time. *As the initial number of infected cells is increased, the time to reach the deterministic threshold decreases. Within this plausible range of 1-1000 initially infected cells, the estimates of the stochastic interval decrease from median 5 to median 1 day. This affects estimations, bounded by this 4 day window. For each value of I(0), 1000 simulations were performed*.

**Supplementary figure 5.**
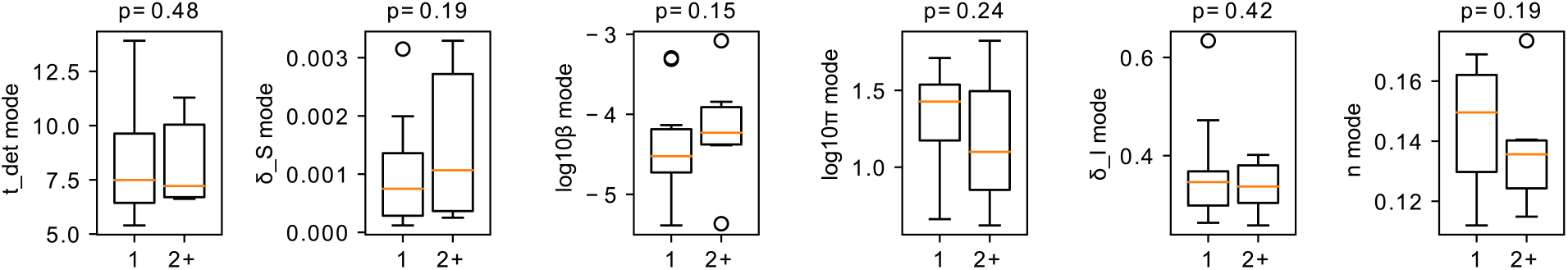
Relationship between viral dynamic parameters and multiple founder status. *Grouped as either 1 founder (n = 24) or 2+ founders (n = 9) No estimated parameter is significantly (Mann-Whitney U-test p-value above each panel) different across founders, meaning the method is not significantly affected by the presence of multiple founders*.

**Supplementary figure 6.**
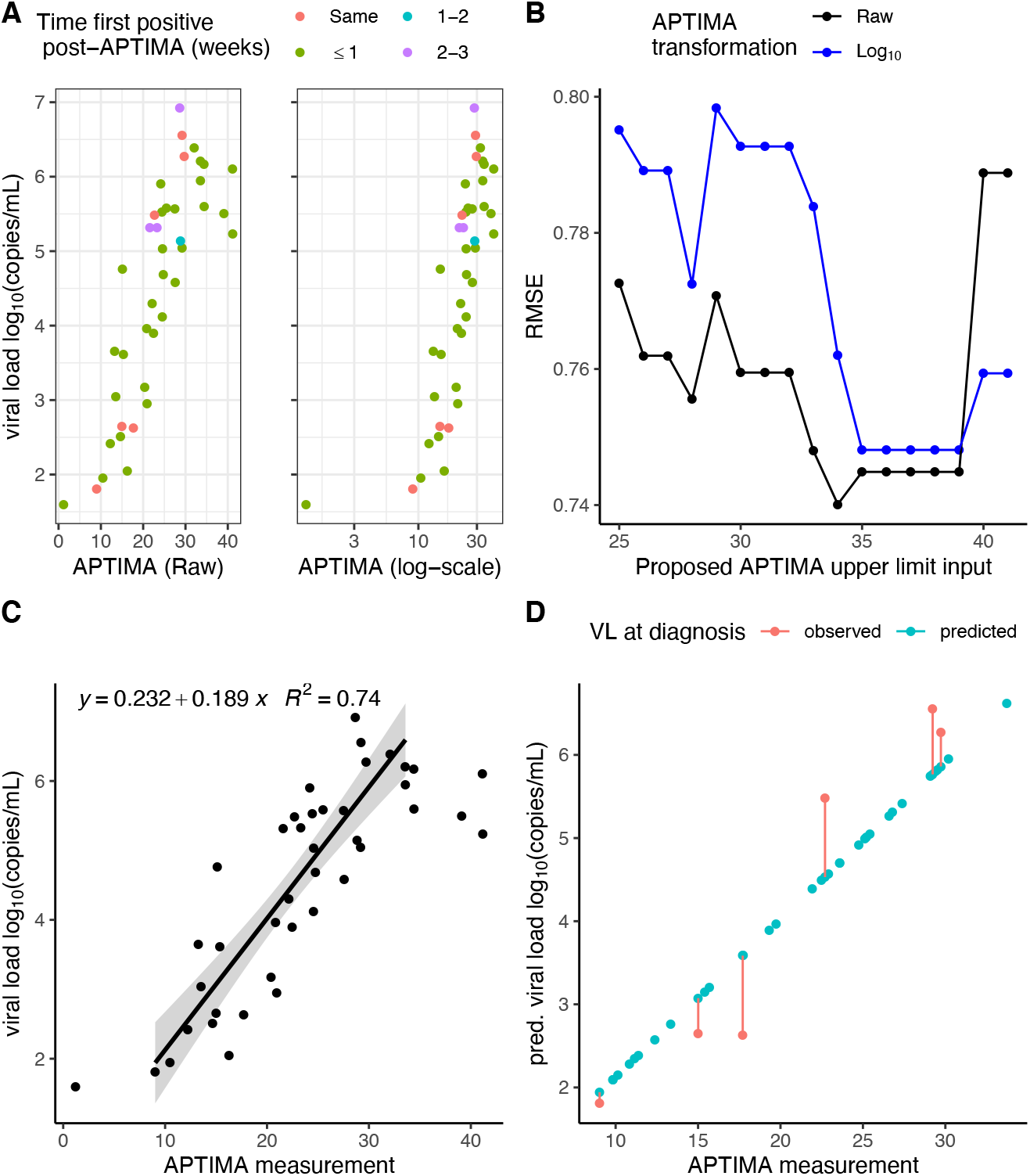
Predicting viral load (VL) with APTIMA measurement at diagnosis. *A) First positive viral load measurements* vs. *concurrent raw or log-transformed APTIMA measurements. Color denotes time (weeks) of first positive relative to the diagnostic APTIMA measurement. ‘Same’ denotes participants who were diagnosed by first positive viral load and positive APTIMA on the same visit. B) Residual mean squared error (RMSE) predicting log_10_ first positive viral load with concurrent APTIMA (raw or log-transformed) for varying input data for different APTIMA upper bounds. C) Selected best regression model from B) denoted by the line with shading for standard error for predicting viral load where APTIMA input range limited to 9-34. Raw data denoted by points. D) Predicted first positive viral load using the model depicted in C) and participants’ APTIMA measurements at diagnosis. Red dots denote the 6 participants where first positive and diagnostic APTIMA were measured together, and red line depicts prediction error. Predicted viral load was only used when for participants without viral load measurements at diagnosis*.

**Supplementary Table 1.**
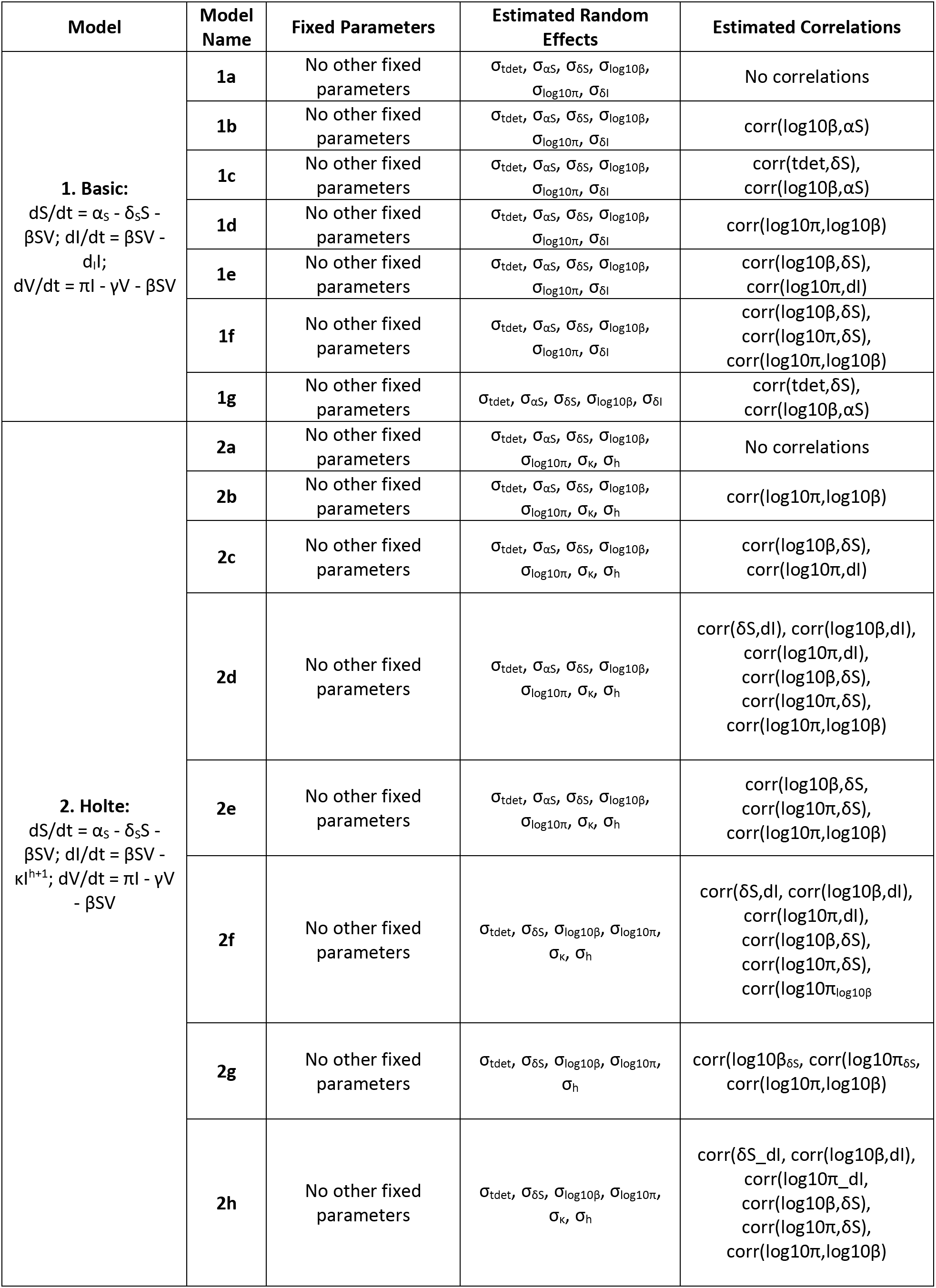
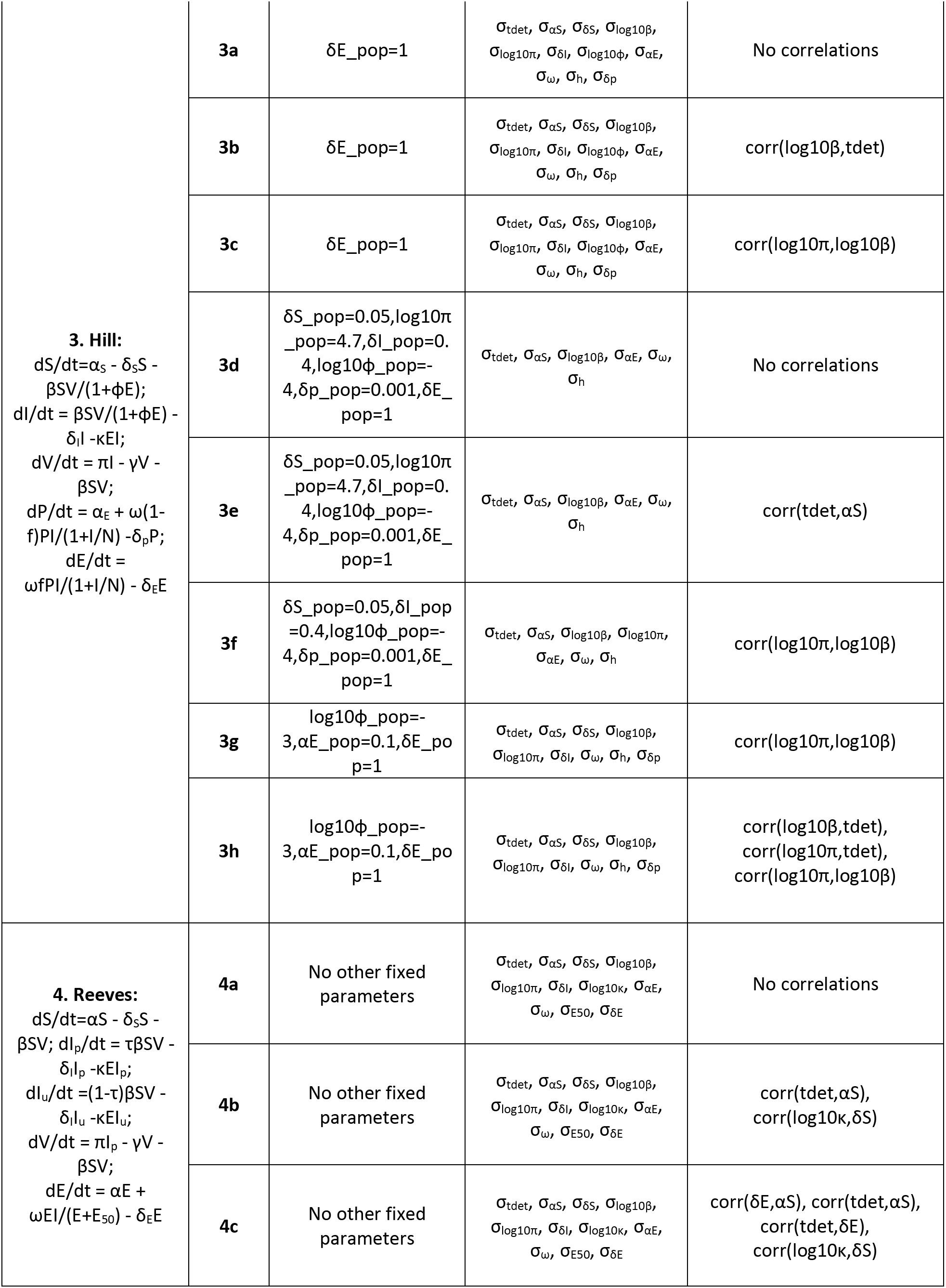
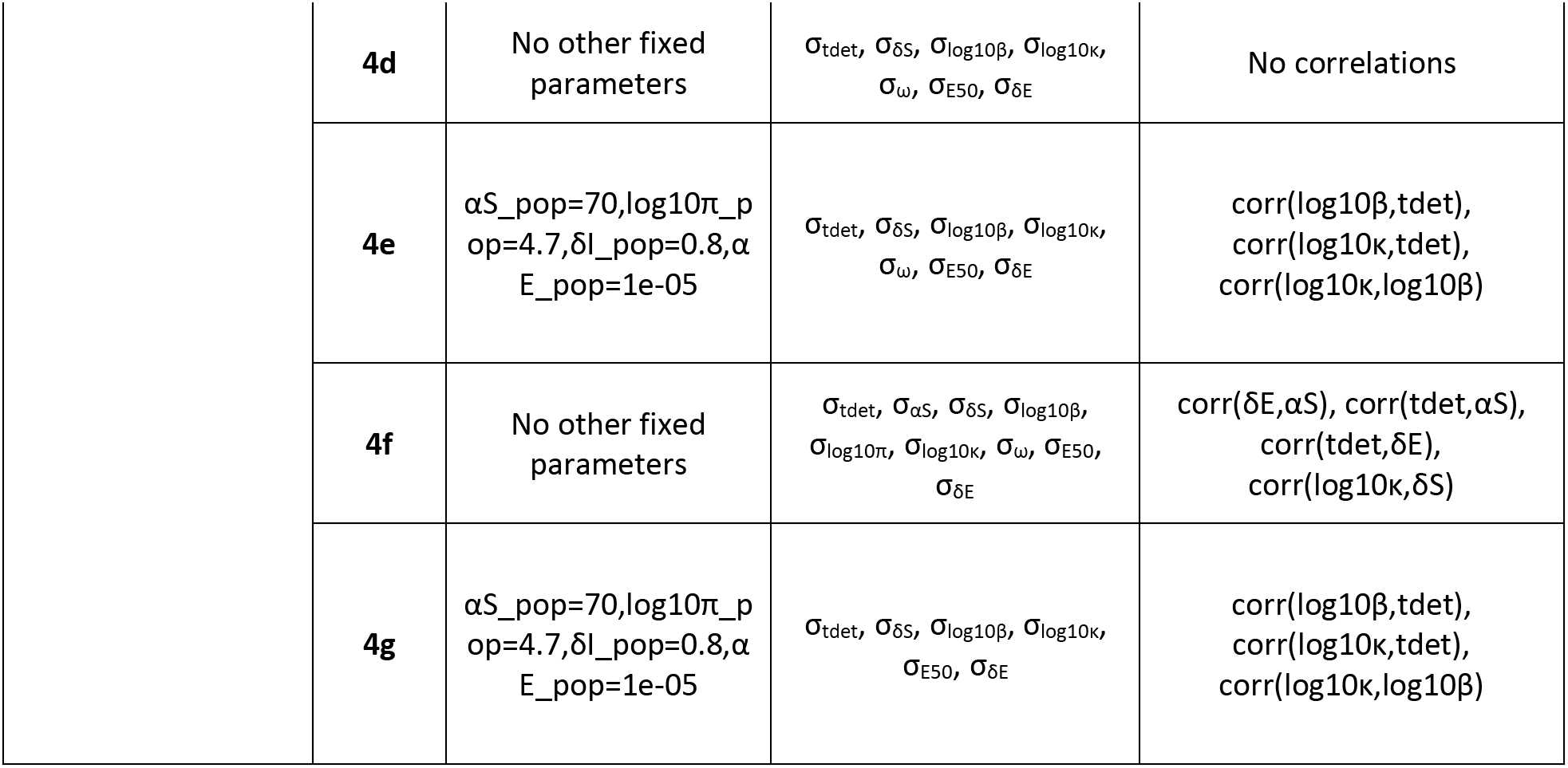
*Description of all the models with their assumptions that were fit to the data. Estimated standard deviation of random effects (a*.*) and correlations for matrix Q for each model are specified, if not included in the table they were assumed to be zero. Other fixed parameters not specified in the text are included here. Distribution for the Log likelihood estimations for each model, AIC for the median and highest likelihood are presented in* ***Supplementary figure 1***.

**Supplementary Table 2.** *6 estimated parameter estimates for each individual. Fixed parameters include V0=0.01 copies/mL and y=23 day^-1^. Parameter α_S_ was assumed identical for all individuals with estimated value 42.7 cells day^-1^μL^-1^*.

| id | t_det | $\delta S$ | $\log 10 \beta$ | $\log 10 \pi$ | $\kappa$ | $\eta$ |
| --- | --- | --- | --- | --- | --- | --- |
| 10066 | 9.62324 | 0.00015132 | -4.65193 | 0.909672 | 0.277224 | 0.123357 |
| 10203 | 8.54516 | 0.00204632 | -3.26718 | 0.817297 | 0.527654 | 0.127292 |
| 10220 | 6.65122 | 0.00313841 | -3.08099 | 0.620871 | 0.401491 | 0.114859 |
| 10428 | 11.1077 | 0.00073429 | -4.59846 | 1.54429 | 0.368804 | 0.111938 |
| 10435 | 19.0919 | 0.00027166 | -4.47328 | 0.904101 | 0.264548 | 0.110447 |
| 10463 | 6.84935 | 0.00026507 | -4.38649 | 0.889934 | 0.301887 | 0.131843 |
| 10723 | 9.97963 | 0.00146281 | -3.99989 | 1.37432 | 0.498574 | 0.141124 |
| 10739 | 9.411 | 0.00019097 | -4.95646 | 1.24387 | 0.261739 | 0.127887 |
| 10742 | 10.3869 | 0.00082387 | -5.0462 | 2.02491 | 0.37825 | 0.1278 |
| 40007 | 6.24852 | 0.00018223 | -5.21088 | 1.54045 | 0.294554 | 0.163842 |
| 40061 | 5.39621 | 0.00314832 | -3.31239 | 0.903244 | 0.445166 | 0.141676 |
| 40094 | 12.6378 | 0.00199422 | -4.13477 | 1.32797 | 0.309711 | 0.13599 |
| 40100 | 11.2924 | 0.00025118 | -5.36688 | 1.82283 | 0.308901 | 0.140513 |
| 40123 | 7.57667 | 0.00329133 | -3.84293 | 1.30979 | 0.365231 | 0.121762 |
| 40168 | 13.9136 | 0.00060717 | -4.75154 | 1.43615 | 0.265229 | 0.168813 |
| 40231 | 9.42558 | 0.00076336 | -4.20229 | 1.12247 | 0.335064 | 0.127668 |
| 40250 | 7.36099 | 0.00109501 | -4.47884 | 1.52832 | 0.348449 | 0.147586 |
| 40257 | 6.78964 | 0.00011896 | -5.1431 | 1.42538 | 0.343796 | 0.168909 |
| 40265 | 6.95792 | 0.00136279 | -4.2837 | 1.42853 | 0.357415 | 0.156733 |
| 40353 | 6.31939 | 0.00143253 | -3.2951 | 0.661891 | 0.47208 | 0.115288 |
| 40363 | 6.62144 | 0.00146629 | -4.34715 | 1.55672 | 0.385113 | 0.139406 |
| 40511 | 5.98824 | 0.00059542 | -4.18057 | 1.39129 | 0.633979 | 0.165386 |
| 40512 | 7.61647 | 0.00135145 | -4.56775 | 1.71063 | 0.365675 | 0.151776 |
| 40577 | 9.63581 | 0.00013342 | -5.38839 | 1.54198 | 0.263169 | 0.151545 |
| 10753 | 6.92639 | 0.00026048 | -4.84951 | 1.30014 | 0.288365 | 0.179543 |
| 40032 | 9.56705 | 0.00444984 | -4.01053 | 1.99893 | 0.763984 | 0.143641 |
| 40211 | 13.6817 | 0.00058183 | -4.29235 | 1.307 | 0.462561 | 0.173981 |
| 40503 | 6.45906 | 0.00070623 | -4.67312 | 1.57485 | 0.35149 | 0.156667 |
| 40646 | 5.3783 | 0.00013301 | -4.82366 | 1.06741 | 0.294896 | 0.131254 |
| 10204 | 17.0122 | 0.00036347 | -4.53213 | 1.08932 | 0.280075 | 0.116799 |
| 10374 | 10.5916 | 0.00273136 | -3.58615 | 1.49338 | 0.847886 | 0.21121 |
| 40067 | 16.4039 | 0.00214128 | -4.33294 | 1.95401 | 0.629711 | 0.1607 |
| 40096 | 11.6224 | 0.00035585 | -4.74625 | 1.35842 | 0.316595 | 0.145131 |
| 40134 | 13.9472 | 0.00026626 | -4.17889 | 0.794832 | 0.359614 | 0.143679 |
| 40139 | 22.4029 | 0.00150565 | -3.88411 | 1.71941 | 1.07298 | 0.103824 |
| 40242 | 20.7467 | 0.00125992 | -3.90151 | 1.03766 | 0.357604 | 0.126244 |
| 40283 | 13.8411 | 0.000963 | -4.25141 | 1.62302 | 0.640396 | 0.106864 |
| 40195 | 19.2263 | 0.00180051 | -4.28104 | 1.76886 | 0.550271 | 0.169485 |
| 40435 | 18.0127 | 0.00076233 | -4.39397 | 1.53545 | 0.497526 | 0.190491 |
| 40492 | 12.2856 | 0.00071631 | -4.55015 | 1.37054 | 0.300422 | 0.199821 |
| 40528 | 2.51471 | 0.011889 | -3.0486 | 2.29511 | 3.53815 | 0.213035 |
| 40652 | 15.3836 | 0.00036841 | -3.87475 | 0.703953 | 0.421935 | 0.173104 |
| 40700 | 24.9363 | 0.00383498 | -4.09306 | 1.66294 | 0.407487 | 0.160303 |
| 40436 | 10.8677 | 0.00066651 | -4.11294 | 0.839844 | 0.257823 | 0.173444 |
| 40491 | 13.3018 | 0.00023218 | -4.43882 | 0.892541 | 0.299901 | 0.142118 |
| 40737 | 32.6352 | 0.00028113 | -4.51066 | 0.978777 | 0.27742 | 0.128034 |

**Supplementary Table 3.** *Best estimate of infection time relative to first positive viral load for each individual. This table illustrates 5 previously reported methods*^3^ *and our population nonlinear mixed effects (pNLME) viral dynamics modeling approach. These methods are the maximum slope of any two points on the upslope (max_slope), the best log-linear regression slope (linear_model), self-reported entries from trial participants (diary), Bayesian phylogenetic inference of median time to most-recent common ancestor (BEAST), and Poisson fitter diversity estimate assuming star-like phylogeny*.

| id | max-slope | linear_model | Diary | BEAST | PFitter | pNLME |
| --- | --- | --- | --- | --- | --- | --- |
| 10066 | -7.0240004 | -18.176592 |  | -2.62 | -8 | -16.372267 |
| 10203 | -10.247329 | -25.198679 |  |  |  | -13.74697 |
| 10220 | 1.18944995 | -2.6721271 |  |  | -13 | -16.04084 |
| 10428 | -18.571441 | -19.277159 |  | -12.18 | -17 | -14.198643 |
| 10435 | -27.053564 | -92.332248 |  |  |  | -26.6943 |
| 10463 | -4.1810206 | -7.290377 |  | -25.42 | -7 | -12.977523 |
| 20225 | -34.189189 | -34.189189 |  | -4.56 | -6 |  |
| 20245 |  |  |  |  |  |  |
| 20263 | -4.3846261 | -7.4981795 |  |  |  |  |
| 20314 | -20.822782 | -23.49317 |  | -3.25 | -10 |  |
| 20337 | -5.4536831 | -11.143324 |  |  |  |  |
| 20355 | -10.400144 | -22.516374 |  |  |  |  |
| 20368 | -3.07086 | -7.8134674 |  | 1.87 | -2 |  |
| 20442 | -5.2628089 | -8.7491781 |  |  |  |  |
| 20502 | -7.7774033 | -13.562055 |  |  | -4 |  |
| 20507 | -5.4597332 | -9.8952878 |  | -9.64 | -7 |  |
| 20509 | -8.4397215 | -17.466691 |  | -8.9 | -6 |  |
| 20511 | -15.132734 | -31.487589 |  | -0.9 | -2 |  |
| 20631 | -12.120706 | -12.731752 |  | -11.29 | -30 |  |
| **30112** | -3.4486448 | -10.198418 |  | -10.97 | -18 |  |
| **30124** | -6.9575257 | -15.019169 |  |  | -16 |  |
| **30190** | -6.0162679 | -19.733117 |  | -1.35 | -6 |  |
| **30507** | -13.070499 | -21.902597 |  |  |  |  |
| **30812** | -5.3750022 | -10.439993 |  |  |  |  |
| **30924** | -26.677419 | -26.677419 |  | -7.82 | -11 |  |
| **40007** | -4.0959219 | -8.7359897 |  | -6.17 | -4 | -10.715117 |
| **40061** | -1.3961514 | -4.0645918 | -13 | 0.39 | -7 | -13.25748 |
| **40094** | -9.3396373 | -28.128959 |  | -19.12 | -20 | -22.087233 |
| **40100** | -15.397661 | -16.900742 | -8 | 1.37 | 2 | -15.072733 |
| **40123** | -2.5469005 | -5.6085575 | -7 |  | -36 | -15.28629 |
| **40168** | -0.3014733 | -2.8255718 | -4 | -6.87 | -6 | -20.232067 |
| **40231** | -9.2630352 | -18.909093 | -7 | -6.45 | -9 | -14.589803 |
| **40250** | -5.1308885 | -10.832909 | -15 | -5.69 | -2 | -12.941387 |
| **40257** | -3.714285 | -6.8801494 | -9 | -6.1 | -11 | -9.51851 |
| **40265** | -3.6988264 | -6.4509726 | -8 | -21.74 | -19 | -13.166543 |
| **40353** | -0.8379281 | -3.7858275 | -3 | -0.46 | 6 | -13.518597 |
| **40363** | -3.4435622 | -7.4401583 | -9 |  | -2 | -11.435493 |
| **40436** | -6.498836 | -19.142592 | -5 | 1.14 | -6 | -18.718233 |
| **40491** | -14.437486 | -28.648138 | -14 |  |  | -17.9225 |
| **40511** |  |  | -3 | -5.65 | -18 | -8.7821767 |
| **40512** | -5.2047177 | -11.910279 | -20 | 2.62 | 2 | -12.201273 |
| **40577** | -7.5341659 | -16.900867 |  | -6.39 | -14 | -14.813207 |

## Notes

### Competing Interest Statement

The authors have declared no competing interest.

### Funding Statement

DBR is funded by a Washington Research Foundation postdoctoral fellowship. Funding was also provided by the National Institute of Health NIAID (K25 AI155224 to DBR, UM1 AI068635 to PBG and JTS, R01 AI150500 to EFC and JTS. The funders had no role in study design, data collection and analysis, decision to publish, or preparation of the manuscript.

### Author Declarations

RV217

### Summary of Updates

Added some funding information and fixed a middle initial.

## References

1 Giorgi EE, Funkhouser B, Athreya G, Perelson AS, Korber BT, Bhattacharya T. Estimating time since infection in early homogeneous HIV-1 samples using a poisson model. BMC Bioinformatics 2010;11:532. https://doi.org/10.1186/1471-2105-11-532.

2 Puller V, Neher R, Albert J. Estimating time of HIV-1 infection from next-generation sequence diversity. PLoS Comput Biol 2017;13:1–20. https://doi.org/10.1371/journal.pcbi.1005775.

3 Rolland M, Tovanabutra S, Dearlove B, Li Y, Owen CL, Lewitus E, et al. Molecular dating and viral load growth rates suggested that the eclipse phase lasted about a week in HIV-1 infected adults in East Africa and Thailand. PLOS Pathog 2020;16:e1008179. https://doi.org/10.1371/journal.ppat.1008179.

4 Fiebig EW, Busch MP, Wright DJ, Kleinman SH, Rawal BD, Garrett PE, et al. Dynamics of HIV viremia and antibody seroconversion in plasma donors: Implications for diagnosis and staging of primary HIV infection. Aids 2003;17:1871–9. https://doi.org/10.1097/01.aids.0000076308.76477.b8.

5 Delaney KP, Hanson DL, Masciotra S, Ethridge SF, Wesolowski L, Owen SM. Time Until Emergence of HIV Test Reactivity Following Infection With HIV-1: Implications for Interpreting Test Results and Retesting After Exposure 2017;64:53–9. https://doi.org/10.1093/cid/ciw666.

6 Pilcher CD, Porco TC, Facente SN, Grebe E, Delaney KP, Masciotra S, et al. A generalizable method for estimating duration of HIV infections using clinical testing history and HIV test results. AIDS 2019;33:1231–40. https://doi.org/10.1097/QAD.0000000000002190.

7 Rossenkhan R, Rolland M, Labuschagne JPL, Ferreira R, Magaret CA, Carpp LN, et al. Combining Viral Genetics and Statistical Modeling to Improve HIV-1 Time-of-infection Estimation towards Enhanced Vaccine E ffi cacy Assessment 2019.

8 Robb ML, Eller LA, Kibuuka H, Rono K, Maganga L, Nitayaphan S, et al. Prospective Study of Acute HIV-1 Infection in Adults in East Africa and Thailand. N Engl J Med 2016:NEJMoa1508952. https://doi.org/10.1056/NEJMoa1508952.

9 Cohen MS, Shaw GM, McMichael AJ, Haynes BF. Acute HIV-1 Infection. N Engl J Med 2011;364:1943–54. https://doi.org/10.1056/NEJMra1011874.

10 Konrad BP, Taylor D, Conway JM, Ogilvie GS, Coombs D. On the duration of the period between exposure to HIV and detectable infection. Epidemics 2017;20:73–83. https://doi.org/10.1016Zj.epidem.2017.03.002.

11 Conway JM, Konrad BP, Coombs D. Stochastic Analysis of Pre- and Postexposure Prophylaxis against HIV Infection. SIAM J Appl Math 2013;73:904–28. https://doi.org/10.1137/120876800.

12 Reeves DB, Huang Y, Duke ER, Mayer BT, Fabian Cardozo-Ojeda E, Boshier FA, et al. Mathematical modeling to reveal breakthrough mechanisms in the HIV Antibody Mediated Prevention (AMP) trials. PLoS Comput Biol 2020;16:1–27. https://doi.org/10.1371/journal.pcbi.1007626.

13 Liu J, Ghneim K, Sok D, Bosche WJ, Li Y, Chipriano E, et al. Antibody-mediated protection against SHIV challenge includes systemic clearance of distal virus. Science (80-) 2016;353:1045–9. https://doi.org/10.1126/science.aag0491.

14 Hessell AJ, Poignard P, Hunter M, Hangartner L, Tehrani DM, Bleeker WK, et al. Effective, low-titer antibody protection against low-dose repeated mucosal SHIV challenge in macaques. Nat Med 2009;15:951–4. https://doi.org/10.1038/nm.1974.

15 Perelson AS, Ribeiro RM. Modeling the within-host dynamics of HIV infection. BMC Biol 2013;11:96. https://doi.org/10.1186/1741-7007-11-96.

16 Reeves DB, Peterson CW, Kiem H, Schiffer JT. Autologous Stem Cell Transplantation Disrupts Adaptive Immune Responses during Rebound Simian/Human Immunodeficiency Virus Viremia. J Virol 2017;91:e00095--17. https://doi.org/10.1128/JVI.00095-17.

17 Borducchi EN, Liu J, Nkolola JP, Cadena AM, Yu W-H, Fischinger S, et al. Antibody and TLR7 agonist delay viral rebound in SHIV-infected monkeys. Nature 2018. https://doi.org/10.1038/s41586-018-0600-6.

18 Holte SE, Melvin AJ, Mullins JI, Tobin NH, Frenkel LM. Density-dependent decay in HIV-1 dynamics. JAIDS J Acquir Immune Defic Syndr 2006;41:266–76. https://doi.org/10.1097/01.qai.0000199233.69457.e4.

19 Davenport MP, Zhang L, Shiver JW, Casmiro DR, Ribeiro RM, Perelson AS. Influence of peak viral load on the extent of CD4+ T-cell depletion in simian HIV infection. J Acquir Immune Defic Syndr 2006;41:259–65. https://doi.org/10.1097/01.qai.0000199232.31340.d3Vi00126334-200603000-00001 [pii].

20 Robb ML, Eller LA, Kibuuka H, Rono K, Maganga L, Nitayaphan S, et al. Prospective Study of Acute HIV-1 Infection in Adults in East Africa and Thailand. N Engl J Med 2016:NEJMoa1508952. https://doi.org/10.1056/NEJMoa1508952.

21 Mayer BT, Krantz EM, Swan D, Ferrenberg J, Simmons K, Selke S, et al. Transient Oral Human Cytomegalovirus Infections Indicate Inefficient Viral Spread from Very Few Initially Infected Cells. J Virol 2017;91:2701–13. https://doi.org/10.1128/JVI.00380-17.

22 Mayer BT, Matrajt L, Casper C, Krantz EM, Corey L, Wald A, et al. Dynamics of persistent oral cytomegalovirus shedding during primary infection in ugandan infants. J Infect Dis 2016;214:1735–43. https://doi.org/10.1093/infdis/jiw442.

23 Ganusov V V, De Boer RJ. Do most lymphocytes in humans really reside in the gut? Trends Immunol 2007;28:514–8. https://doi.org/10.1016/j.it.2007.08.009.

24 Gillespie DT. Approximate accelerated stochastic simulation of chemically reacting systems. J Chem Phys 2001;115:1716–33. https://doi.org/10.1063/L1378322.

25 Hughes JP, Baeten JM, Lingappa JR, Magaret AS, Wald A, de Bruyn G, et al. Determinants of Per-Coital-Act HIV-1 Infectivity Among African HIV-1-Serodiscordant Couples. J Infect Dis 2012;205:358–65. https://doi.org/10.1093/infdis/jir747.

26 Drummond AJ, Suchard MA, Xie D, Rambaut A. Bayesian phylogenetics with BEAUti and the BEAST 1.7. Mol Biol Evol 2012;29:1969–73. https://doi.org/10.1093/molbev/mss075.

27 Giorgi EE, Funkhouser B, Athreya G, Perelson AS, Korber BT, Bhattacharya T. Estimating time since infection in early homogeneous HIV-1 samples using a poisson model. BMC Bioinformatics 2010;11:532. https://doi.org/10.1186/1471-2105-11-532.

28 Janes H, Herbeck JT, Tovanabutra S, Thomas R, Frahm N, Duerr A, et al. HIV-1 infections with multiple founders are associated with higher viral loads than infections with single founders. Nat Med 2015;21:1139–41. https://doi.org/10.1038/nm.3932.

29 Gilbert PB, Juraska M, DeCamp AC, Karuna S, Edupuganti S, Mgodi N, et al. Basis and Statistical Design of the Passive HIV-1 Antibody Mediated Prevention (AMP) Test-of-Concept Efficacy Trials. Stat Commun Infect Dis 2017;9:. https://doi.org/10.1515/scid-2016-0001.

30 Huang Y, Karuna S, Carpp LN, Reeves D, Pegu A, Seaton K, et al. Modeling cumulative overall prevention efficacy for the VRC01 phase 2b efficacy trials. Hum Vaccines Immunother 2018;0:1-12. https://doi.org/10.1080/21645515.2018.1462640.

31 Ramratnam B, Bonhoeffer S, Binley J, Hurley A, Zhang L, Mittler JE, et al. Rapid production and clearance of HIV-1 and hepatitis C virus assessed by large volume plasma apheresis. Lancet 1999;354:1782–5. https://doi.org/10.1016/S0140-6736(99)02035-8.

32 Ejima K, Kim KS, Ito Y, Iwanami S, Ohashi H, Koizumi Y, et al. Inferring Timing of Infection Using Within-host SARS-CoV-2 Infection Dynamics Model: Are ‘Imported Cases’ Truly Imported? *MedRxiv* 2020;4297:2020.03.30.20040519. https://doi.org/10.1101/2020.03.30.20040519.

33 Beier P, Burnham KP, Anderson DR. Model Selection and Inference: A Practical Information-Theoretic Approach. vol. 65. 2001.

